# Inhibition of SHP2 ameliorates psoriasis by decreasing TLR7 endosome localization

**DOI:** 10.1101/2020.09.28.20202861

**Authors:** Yuyu Zhu, Fenli Shao, Wei Yan, Zhen Bouman Chen, Bowen Ke, Xian Jiang, Haibo Cheng, Dongdong Sun, Chenglin Song, Lingdong Kong, Wenjie Guo, Yuping Lai, Gen-Sheng Feng, Qiang Xu, Yang Sun

**Author notes:** Corresponding to Yang Sun, Ph.D., Professor or Qiang Xu, Ph.D.,Professor, School of Life Sciences, Nanjing University, Nanjing 210023 China, (Y. Sun) or (Q. Xu). These authors contributed equally to this work (Yuyu Zhu, Fenli Shao, Wei Yan).

## Abstract

Psoriasis is a complex chronic inflammatory skin disease with unclear molecular mechanisms. Here, we identify Src homology-2 domain containing protein tyrosine phosphatase-2 (SHP2) as a novel accelerator of psoriasis development. Both genetic ablation of SHP2 in macrophages and pharmacological inhibition of SHP2 prevents the development of psoriasis-like skin inflammation in an imiquimod-induced murine model of psoriasis. Mechanistically, SHP2 promotes the trafficking of Toll-like receptor 7 (TLR7) from Golgi to endosome through its interaction with and dephosphorylation of TLR7 at Tyr1024, which promotes the ubiquitination of TLR7 and psoriasis-like skin inflammation. Importantly, SHP2 allosteric inhibitor SHP099 reduces the expression of pro-inflammatory cytokines in peripheral blood mononuclear cells from human patients with psoriasis. Collectively, our findings identify SHP2 as a novel regulator of psoriasis and suggest that SHP2 inhibition may be a promising therapeutic approach for psoriatic patients.

## Introduction

Psoriasis is a common and complex chronic inflammatory skin disease that is characterized by epidermal hyperplasia, aberrant differentiation of keratinocytes, exaggerated angiogenesis, and dermal infiltration of inflammatory cells (Boehncke and Schön, 2015; Lebwohl, 2018; Perera et al., 2012). It affects 2-3% of the global population (Crow, 2012), but the exact mechanism underlying its development remains unclear. The imiquimod (IMQ)-induced mouse model largely mimics the phenotype of psoriasis in human (van der Fits et al., 2009) and has been widely used in pre-clinical study of psoriasis. Using the IMQ-induced mouse model, recent studies have identified the involvement of IL-23/IL-17 axis in psoriasis (Boehncke and Schön, 2015; Burkett and Kuchroo, 2016; Kopp et al., 2015; Lebwohl, 2019) Activated macrophages and dendritic cells (DCs) generate IL-23, TNF-α, IL-6, and IL-1β to stimulate and polarize helper T (Th) cells to transition into Th1, Th17, and Th22 cell subsets, which respectively IFN-γ, IL-17A/F, and IL-22. These proinflammatory cytokines then promote the proliferation and activation of keratinocyte cells, which then produce IL-23 and aggravate the inflammation and psoriasis progression. In patients with psoriasis, the major TNF-α-producing cells are the slan^+^ macrophages (Brunner et al., 2013). The reduction of these cells during remission of psoriasis underlines their critical role in disease development, maintenance, or both (Brunner et al., 2013), emphasizing an important role for macrophages in the development of psoriasis.

Other possible effectors of psoriasis are the toll-like receptors (TLRs), which are essential sensors of a variety of viral and microbial infections and endogenously derived damage-associated molecular patterns (DAMPs) (Bryant et al., 2015). TLRs recruit adaptor proteins and activate transcription factors to produce inflammatory cytokines that promote the initiation of adaptive immune responses (Newman et al., 2016). The TLR family consists of 10 members in humans and 13 in mice, which are expressed both on the cell surface and intracellularly (Barbalat et al., 2011). The intracellular TLRs, including TLR3, TLR7, TLR8, and TLR9 in humans, recognize nucleic acids (Blasius and Beutler, 2010). TLR7 recognizes single-stranded RNA and traffics to endosomes (Wang et al., 2020). Imiquimod (IMQ) is a TLR7 ligand and has been used for topical treatment of genital and perianal warts caused by human papilloma virus (Beutner and Tyring, 1997). Interestingly, topical treatment with IMQ in patients of actinic keratoses and superficial basal cell carcinomas can exacerbate psoriasis even if it was previous well controlled (Gilliet et al., 2004). Such IMQ-exacerbated psoriasis occurs not only in the topically treated areas, but also at distant skin sites that were previously unaffected (Wu et al., 2004). These findings suggest that TLR7 activation may promote psoriasis, which has not been identified.

Src homology-2 domain containing protein tyrosine phosphatase-2 (SHP2) is a ubiquitously expressed cytoplasmic tyrosine phosphatase that contains two SH2 domains and one catalytic protein tyrosine phosphatase (PTP) domain (Feng et al., 1993). It is encoded by *PTPN11* and has critical functions in cell proliferation, differentiation, and survival (Qu, 2002). In the basal state, SHP2 is inactive in the basal state, but when combined with a phosphotyrosine (pY) protein, it activates as a phosphatase to elicit downstream signaling (Barford and Neel, 1998). SHP2 missense mutations account for approximately 50% of the cases of Noonan syndrome (Keilhack et al., 2005), a common autosomal dominant disorder characterized by multiple, variably penetrant defects. Intriguingly, a 55-year-old woman with many cardinal physical features of Noonan syndrome also suffered simultaneously from annular pustular psoriasis (Catharino et al., 2016), suggesting a potential role of SHP2 in psoriasis. However, this has not been investigated either *in vitro* or *in vivo*.

In the present study, we report that SHP2 promotes the pathogenesis of psoriasis by enhancing TLR7 trafficking to the endosome. Ablation or inhibition of SHP2 in macrophages weakens NF-κB activation, subsequently resulting in amelioration of IMQ-induced psoriasis-like skin inflammation in mice. Mechanistically, SHP2 dephosphorylates TLR7 at Tyr1024 to promote its ubiquitination, trafficking to the endosome, without affecting TLR7 expression. Collectively, our findings identify SHP2 as a critical regulator of psoriasis and as a potential therapeutic target for the treatment of this skin disease.

## Results

### SHP2 expression is increased in both human psoriatic patients and IMQ-induced psoriasis-like murine model

To assess whether SHP2 may play a functional role in psoriasis, we first analyzed the expression and activity of SHP2 in human samples collected healthy donors (normal controls; Normal) and psoriatic patients. At the mRNA level, SHP2 (encoded by *PTPN11* gene) was significantly higher in skin lesions (Figure 1A) and human peripheral blood mononuclear cells (PBMCs) (Figure 1B) derived from psoriatic patients than those from normal controls. At the protein level, PBMCs (Figure 1C) and skin biopsies (Figure 1D) from psoriatic patients also showed a strong increase in SHP2, when compared to normal controls. Consistently, enzymatic assays revealed an increased SHP2 activity in PBMCs from psoriatic patients vs. normal controls (Figure 1E). Additionally, the level of p-ERK, which has been shown to be positively regulated by SHP2, was also higher in the skin lesions from psoriatic patients than those from normal controls (Figure 1F).

**Figure 1.**
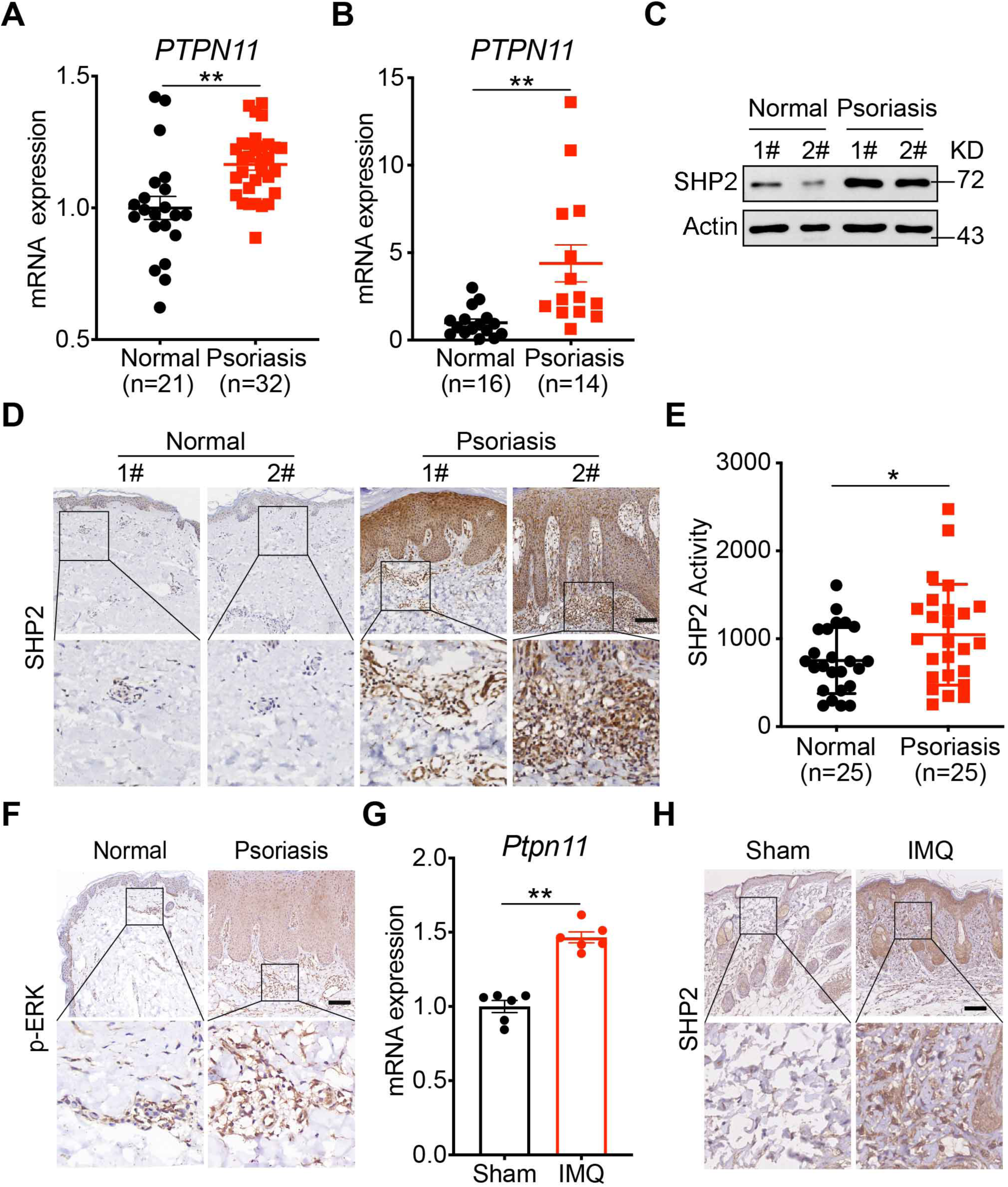
SHP2 expression is increased in both human psoriatic patients and IMQ-induced psoriasis-like murine model. (A) Expression of *PTPN11* (*gene encoding SHP2*) in skin lesions in psoriatic patients compared with skin from healthy donors based on microarray data (No. GSE14905). (B) Expression of *PTPN11* in human PBMCs from psoriatic patients (n=14) and normal controls (n=16). (C) Western blot analysis of PBMCs lysates derived from psoriatic patients and normal controls. (D) Representative SHP2 staining in skin sections from psoriatic patients (n=13) and normal controls (n=5). Scale bars: 200 μm. (E) The catalytic activity of SHP2 was measured in human PBMCs lysates derived from psoriatic patients (n=25) and normal controls (n=25). (F) Representative p-ERK staining of skin sections from psoriatic patients and normal controls. Scale bars: 200 μm. (G) Quantitative PCR analysis of *Ptpn11* mRNA levels in the IMQ-treated or non-treated dorsal back from C57BL/6J mice at day 5 (n=6/group). Data were normalized to GAPDH expression. (H) Representative histological sections of IMQ-treated or non-treated dorsal back from C57BL/6J mice at day 5. Scale bar: 100 μm. Data represent mean ± SEM. *P* values are determined by Two-tailed Mann-Whitney U test (A and B) or Two-tailed Student’s *t* test (E and G). \**P*<0.05, \*\**P*<0.01.

Similarly, SHP2 expression was increased in the dorsal skin of the IMQ-induced psoriasis-like murine model, both at the mRNA and protein levels (Figures 1G and 1H). Taken together, data from human and mouse indicate an increase in SHP2 function in psoriasis.

### SHP2 deficiency in macrophages alleviates the psoriasis-like phenotype in the IMQ-induced murine model

Compared to normal controls, there was a substantial increase in CD68^+^ macrophages infiltration in the skin lesions of psoriatic patients compared to normal controls (Figure 2A). Noticeably, there is a clear colocalization of CD68 and SHP2 in the dermis with psoriasis (Figure 2B). A similar result was also seen in the IMQ-induced psoriasis-like murine model (Figure 2C), suggesting that SHP2 is highly expressed in the infiltrated macrophages in the psoriatic skin.

**Figure 2.**
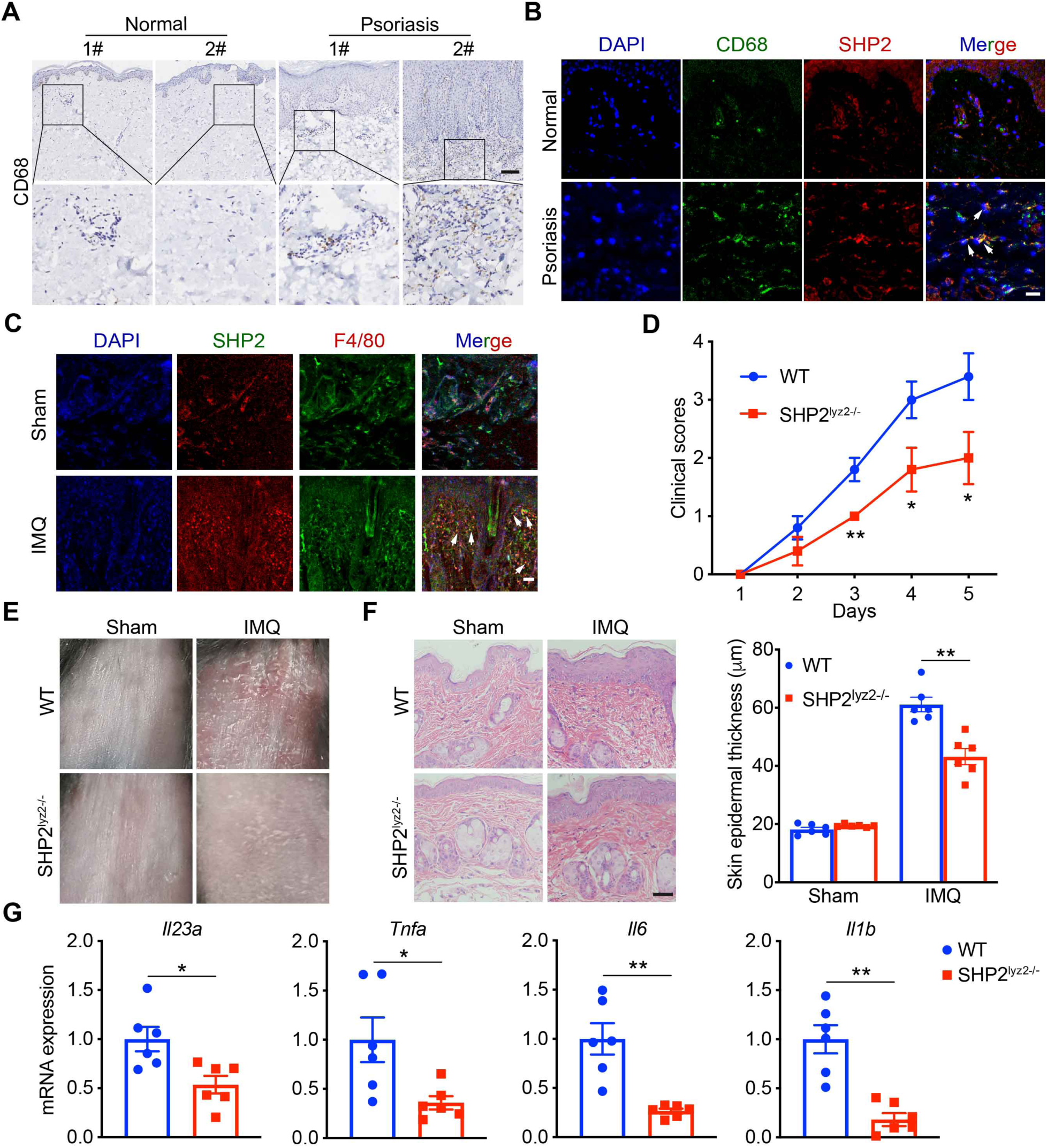
SHP2 deficiency in macrophages alleviates psoriasis-like phenotype in the IMQ-induced murine model. (A) Representative CD68 staining of skin sections from psoriatic patients (n=13) and normal controls (n=5). Scale bar: 200 μm. (B) Immunofluorescence staining of skin sections from psoriatic patients and normal controls with SHP2 and CD68, showing SHP2 expression in macrophages (arrows). SHP2, green; CD68, red. Scale bar: 100 μm. (C) Immunofluorescence staining of skin sections from IMQ-induced and sham mice with SHP2 and F4/80, showing SHP2 expression in macrophages (arrows). SHP2, green; F4/80, red. Scale bar: 100 μm. (D-F) Clinical scores (D), representative images (E) and histological sections of dorsal back (F) from wild-type (n=6) and SHP2^lyz2-/-^ mice (n=6) treated with IMQ for 4 days. Scale bar: 100 μm. Left: H&E (hematoxylin and eosin) staining data; right: statistical data (mean ± SEM). (G) Quantitative PCR analysis of mRNA levels in dorsal skin from wild-type and SHP2^lyz2-/-^ mice. Results were normalized to GAPDH expression and are shown as mean ± SEM. *P* values are determined by two-tailed Student’s *t* test (D, F, G). \**P*<0.05, \*\**P*<0.01.

To determine the causal role of SHP2 in psoriasis, we generated macrophage-specific (Lyz2-Cre) SHP2 conditional knockout mice (SHP2^lyz2-/-^) and treated the animals with IMQ on the dorsal skin. After 4 days of treatment, the wild-type mice exhibited swelling, epidermal acanthosis, and remarkably increased proliferation of keratinocytes and dermal inflammatory cell infiltration. In contrast, all these pathological changes were remarkably ameliorated in SHP2^lyz2-/-^ littermates. revealed a greatly diminished psoriasis pathology in IMQ-treated SHP2^lyz2-/-^ mice (Figures 2D-2F). In line with this phenotype, the skin from SHP2^lyz2-/-^ mice also expressed significantly lower levels of IMQ-activated inflammatory cytokines, i.e. *Il23a, Tnfa, Il6*, and *Il1b* (Figure 2G). Similar decrease in these cytokines were also observed in bone-marrow derived macrophages (BMDMs; Figure S1A) and peritoneal macrophages (PMs; Figure S1B) of SHP2^lyz2-/-^ mice. Notably, such decrease in the pro-inflammatory cytokines in SHP2^lyz2-/-^ mice was recapitulated by SHP2-deficient THP-1 cells, when compared to the control cells (Figure S1C). In addition, when stimulated with IL-36, peritoneal macrophages derived from SHP2^lyz2-/-^ mice also produced less psoriasis-related cytokines than those derived from the wild-type littermates (Figure S2).

To test the possible role of SHP2 in dendritic cells in the observed phenotype, we also generated dendritic cells-specific (Itgax-Cre) SHP2 conditional knockout mice (SHP2^Itgax-/-^). Treatment of these mice and their wild-type littermates with IMQ did not result in any significant difference (Figure S3). Taken together, our data demonstrate that SHP2 deficiency in macrophages causes resistance to IMQ-induced psoriasis.

### SHP2 deficiency mitigates psoriasis by suppressing NF-κB activation

RNA sequencing of peritoneal macrophages isolated from wild-type and SHP2^lyz2- /-^ mice and then treated with IMQ for 4 h *in vitro* revealed an enrichment of cytokines related to NF-κB signaling (Figures 3A and 3B). The decrease of several genes in the NF-κB signaling in macrophages from SHP2^lyz2-/-^ mice was confirmed by quantitative PCR (qPCR) (Figure 3C). Comparison between IMQ-stimulated BMDMs and PMs from wild-type vs. SHP2^lyz2-/-^ mice also demonstrated significantly lower phosphorylation levels of IKKα/β and its downstream p65 in macrophages from SHP2^lyz2-/-^ mice, specifically at 15-60 min upon stimulation (Figure 3D). In parallel, analysis of the skin lesions collected from IMQ-induced mice (Figure 3E) and psoriatic patients (Figure 3F) also revealed significantly higher levels of p-p65 protein in the infiltrated cells. Collectively, these data suggest that knockout of SHP2 in macrophages attenuated the disease severity through attenuating NF-κB activation.

**Figure 3.**
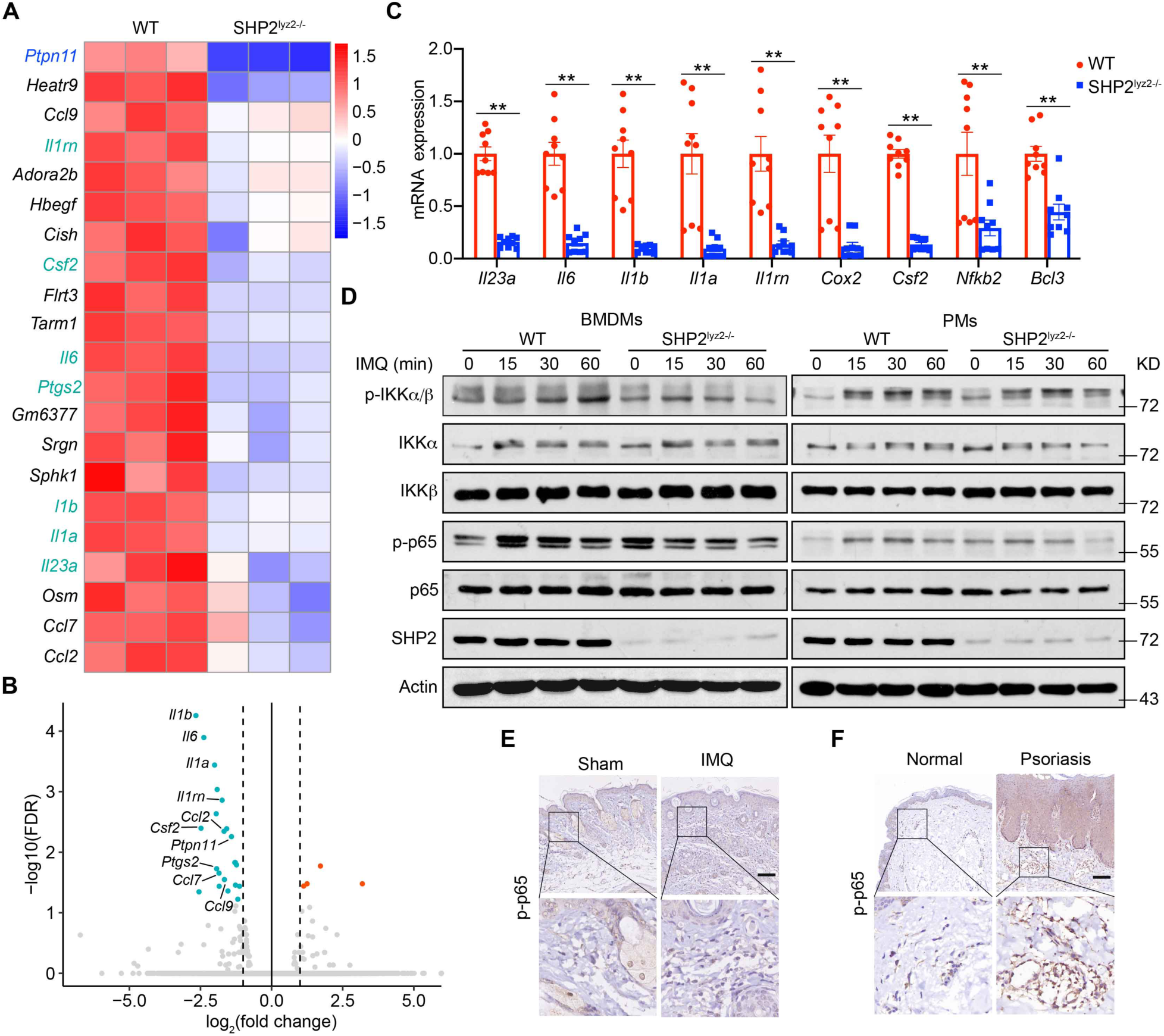
SHP2 deficiency mitigates psoriasis by suppressing NF-κB activation. (A) Heat map showing mRNA expression in peritoneal macrophages derived from wild-type and SHP2^lyz2-/-^ mice after IMQ treatment based on RNA sequencing (n=3/group). Colors represent high (red) and low (blue) intensity. Genes labeled in green indicate they are in the NF-κB signaling. (B) Volcano plot image of up- (in red) and down- (in green) regulated genes in peritoneal macrophages from wild-type and SHP2^lyz2-/-^ mice after IMQ treatment (n=3/group). (C) Expression levels of representative psoriasis-related genes decreased in the IMQ model of SHP2^lyz2-/-^ mice compared to wild-type mice. (D) BMDMs and PMs derived from wild-type and SHP2^lyz2-/-^ mice were stimulated by IMQ (10 µg/ml) for indicated times. Whole cell lysates were subjected to Western blotting. (E and F) Representative p-p65 staining of skin sections from mouse (E) and human (F). Scale bar: 100 μm (E), 200 μm (F). Data represent the mean ± SEM. *P* values are determined by two-tailed Student’s *t* test (C). \*\**P*<0.01.

### SHP2 interacts with TLR7

Since TLR7 has been reported to participate in the development of psoriasis (Kim et al., 2018), and TLR7 at the upstream of the NF-κB signaling pathway, we examined the relationship between SHP2 and TLR7. Next, we performed co-immunoprecipitation (co-IP), which confirms that TLR7 indeed interacted with SHP2, in HEK293T cells overexpressing SHP2 (Figure 4A) or TLR7 (Figure 4B). Immunoprecipitation and immunoblotting of HA or GFP epitope tags from lysates of IMQ-stimulated cells overexpressing HA-tagged SHP2 and GFP-tagged TLR7 revealed a strongly increased interaction between TLR7 and SHP2 (Figures 4C-4E). The endogenous interaction between TLR7 and SHP2 was also augmented in HEK293T cells after IMQ infection (Figure 4F). In agreement with the co-IP results, confocal microscopy also visualized a significant increase in the co-localization of TLR7 and SHP2 in THP-1 cells following IMQ treatment (Figure 4G).

**Figure 4.**
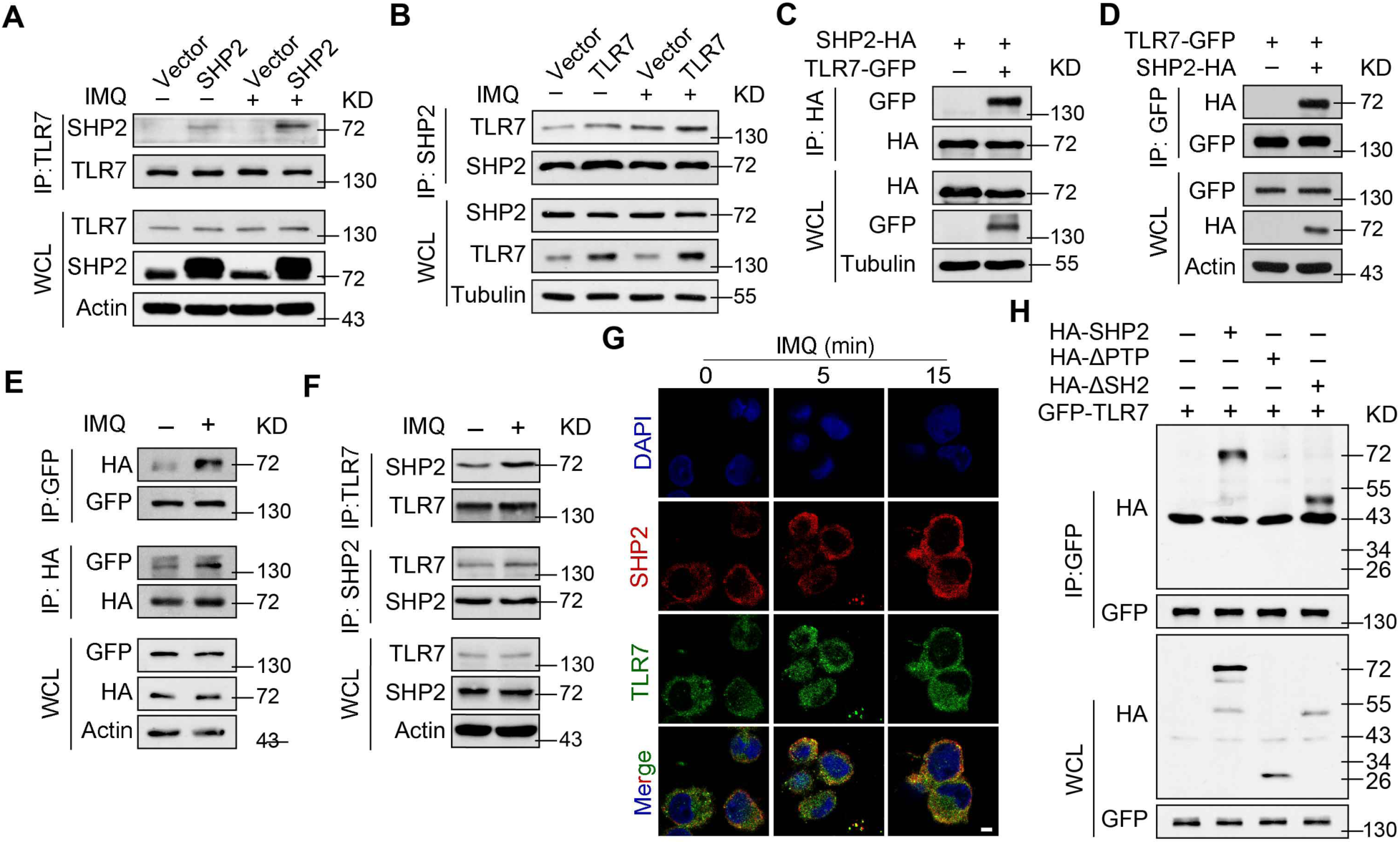
SHP2 interacts with TLR7. (A-B) HEK293T cells were transiently transfected by plasmids expressing SHP2 (A), or TLR7 (B). After 48 h, cells were left unstimulated or stimulated by IMQ (10 µg/ml) for 30 min. Cell lysates underwent immunoprecipitation with TLR7 and SHP2 antibodies and the immunoblotting with indicated antibodies. (C-D) Immunoblotting of reciprocal Co-IP from HEK293T cells overexpressing HA-tagged SHP2 or GFP-tagged TLR7, along with plasmids expressing GFP-tagged TLR7 (C) and HA-tagged SHP2 (D), respectively. After 48 h, cell lysates were immunoprecipitated by anti-HA or anti-GFP and probed by indicated antibodies. (E) HEK293T cells were transfected with plasmids expressing HA-tagged SHP2 and GFP-tagged TLR7. After 48 h, cells were left unstimulated or stimulated by IMQ (10 µg/ml) for 30 min. Cell lysates were immunoprecipitated by anti-HA or anti-GFP and probed by indicated antibodies. (F) HEK293T cells were unstimulated or stimulated by IMQ (10 µg/ml) for 30 min. Cell lysates were immunoprecipitated by anti-TLR7 or anti-SHP2 and probed by indicated antibodies. (G) Representative images of confocal microscopy performed with PMA-differentiated THP-1 cells treated with IMQ (10 µg/ml) for indicated durations. Scale bar: 20 µm. (H) Immunoblotting of HEK293T cells co-transfected for 48 h with GFP-TLR7, plus HA-SHP2, HA-SHP2 mutant vectors, followed by immunoprecipitation with anti-GFP beads.

SHP2 possesses two SH2 domains at the N-terminus, a PTP domain, and a phosphotyrosine-containing tail. To determine the structural basis for its scaffolding function, we generated a series of SHP2 mutants, with PTP or SH2 domain deleted. Deletion of the PTP domain almost completely abolished the association of SHP2 and TLR7 (Figure 4H), suggesting the PTP domain of SHP2 interacts with TLR7.

### SHP2 promotes TLR7 trafficking to endosomes

Next, we interrogated the molecular effect of SHP2 interaction with TLR7.We first found that neither knock down nor over-expression of SHP2 altered the expression of TLR7 in THP-1 cells (Figure 5A; Figure S4A). Likewise, IMQ treatment did not significantly change the TLR7 expression, although it significantly increased the phosphorylation levels of p65 in a time-dependent manner (Figures S4B and S4*C*). However, in the same time course, we observed an accumulation of TLR7 on the cell membrane, which was more obvious in cells overexpressing SHP2 and eliminated when SHP2 was knocked down (Figure 5B). Resiquimod (R848) is a TLR7/8 agonist that induced the consistent effects with IMQ (Figure S5). Analysis of isolated cytoplasm and cell membranes confirmed the increase in TLR7 on the cell membrane when SHP2 was overexpressed (Figure 5C). Collectively, these data suggest that SHP2 promotes TLR7 localization to the cell membrane, without affecting its expression.

**Figure 5.**
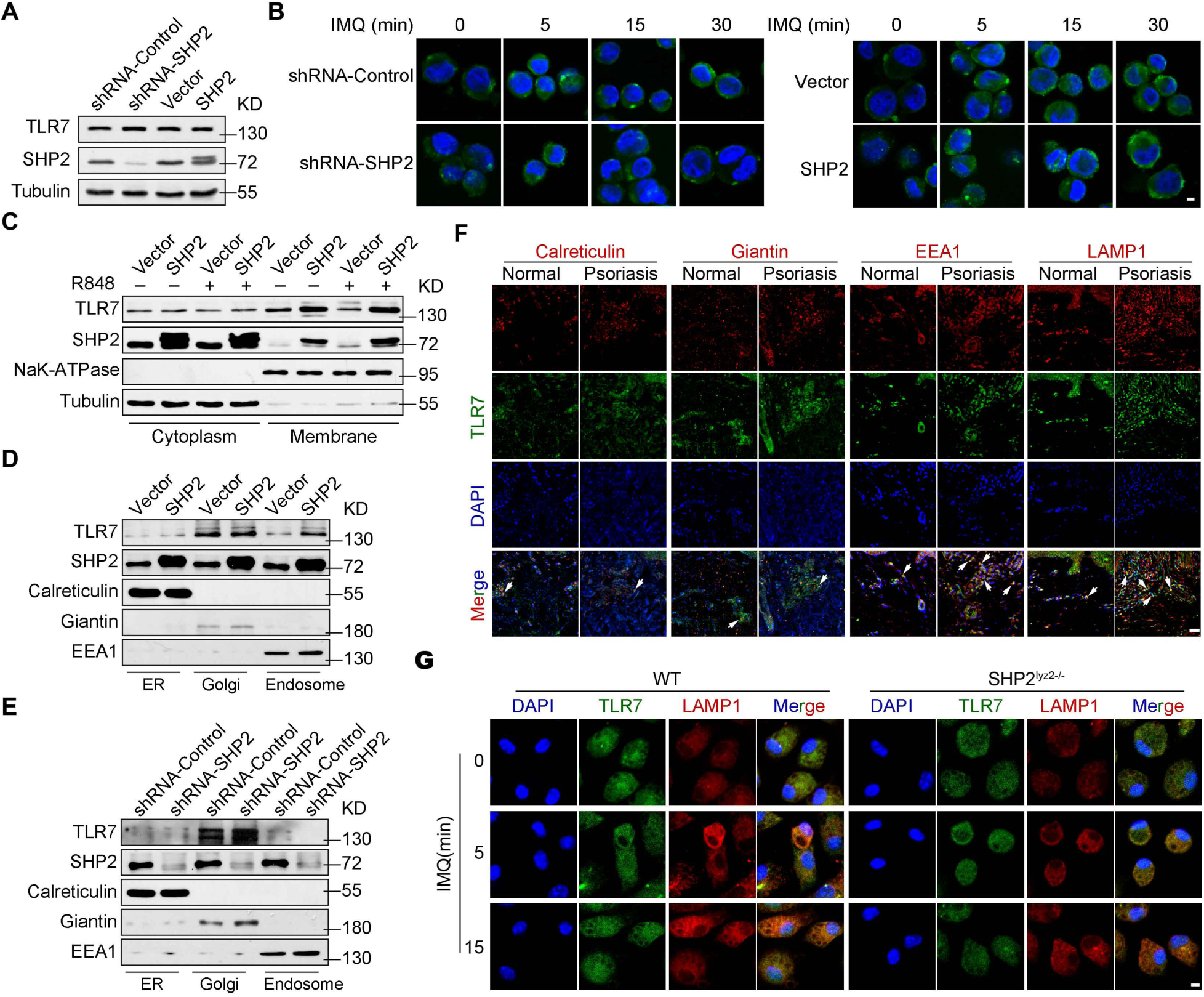
SHP2 promotes TLR7 trafficking to endosomes. (A) Immunoblotting of PMA-differentiated THP-1 cells infected with shRNA-Control, shRNA-SHP2, vector or SHP2 lentiviruses. (B) Confocal microscopy imaging of TLR7 in PMA-differentiated THP-1 cells, lentivirus-infected as indicated, and treated with IMQ (10 µg/ml) for various times. Scale bar: 10 µm. (C) PMA-differentiated THP-1 cells were infected with vector or SHP2 lentivirus and then treated with or without R848 (10 µg/ml). Subcellular fractionation was performed and cytoplasmic and cell membrane proteins were probed by respective antibodies. (D and E) Immunoblot analysis of TLR7 and SHP2 in the isolated ER, Golgi and endosome from PMA-differentiated THP1 cells with either SHP2 overexpressed (D) and SHP2 deficiency(E). (F) Skin sections from psoriatic patients and healthy controls were immunostained for TLR7 together with a marker of the ER (Calreticulin), Golgi (Giantin), early endosome (EEA1) or late endosome/lysosome (LAMP1) prior to analysis by confocal microscopy, showing TLR7 localization in different organelles (arrows). Scale bar: 100 μm. (G) Confocal microscopy imaging of PMs derived from wild-type and SHP2^lyz2-/-^ mice were treated with IMQ (10 µg/ml) for 0-15 minutes and labelled with antibodies against specific proteins. Scale br: 20 µm.

To obtain more details on the TLR7 membrane localization, we isolated endoplasmic reticulum (ER), Golgi, endosomes, and plasma membrane from THP-1 and HEK293T cells. SHP2 overexpression caused an increased level of TLR7 only in the endosomes but not in the ER or Golgi (Figure 5D; Figure S6A). Conversely, in SHP2-deficient cells, TLR7 was decreased in the endosomes (Figure 5E; Figure S6B). Detection of subcellular localization of TLR7 in the human skin revealed comparable levels of TLR7 in the ER and Golgi (marked by calreticulin and giantin, respectively), between psoriatic patients and normal controls (Figure 5F; Figure S7). In contrast, TLR7 was significantly increased in the endosomes of the skin from psoriatic patients, where it was co-located with EEA1 and LAMP1 (Figure 5F). In a complementary experiment, when SHP2 was deleted in PMs, the accumulation of TLR7 in endosome was markedly reduced (Figure 5G; Figure S8). Therefore, data in Figure 5 indicate that SHP2 promotes the trafficking of TLR7 to the endosomes, particularly in the context in psoriasis.

### SHP2 promotes activation of TLR7/NF-κB signaling in a phosphatase-dependent manner

Given that SHP2 is a tyrosine phosphatase, we queried whether SHP2 dephosphorylates tyrosine residues on TLR7. In HEK293T cells co-transfected with GFP-TLR7 and HA-SHP2 (wild-type, WT) vectors and then stimulated by IMQ, we detected phosphor-tyrosine (p-Tyr) in TLR7 pulled down by anti-GFP (Figure 6A, left lane). When the WT SHP2 was replaced with the gain-of-function mutant, i.e. SHP2^D61A^, p-Tyr of TLR7 was inhibited (Figure 6A, middle lane). On the other hand, loss-of-function mutant SHP2^C459S^ intensified p-Tyr of TLR7 (Figure 6A, right lane).

**Figure 6.**
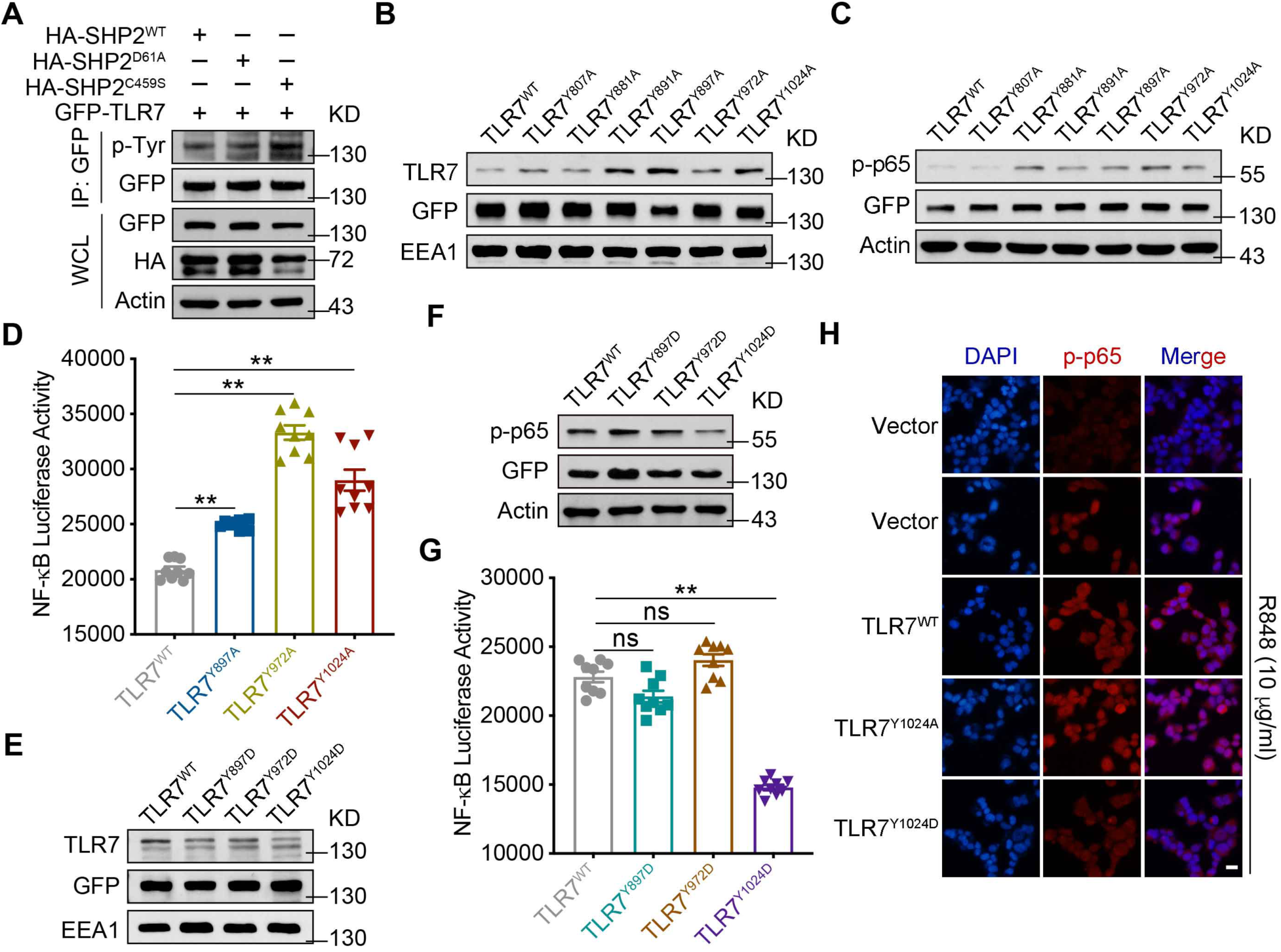
SHP2 promotes activation of TLR7/NF-κB signaling in a phosphatase-dependent manner. (A) Immunoblotting of HEK293T cells co-transfected for 48 h with GFP-TLR7, plus HA-tagged SHP2 or SHP2 mutant vectors, followed by immunoprecipitation with anti-GFP. (B and E) Immunoblotting of TLR7 in the endosome isolated from HEK293T cells that were transfected with GFP-TLR7 or GFP-TLR7 mutant vectors. (C and F) Immunoblotting of p-p65 in lysates of HEK293T cells transfected with GFP-TLR7 and TLR7 mutant vectors and then treated with IMQ (10 µg/ml) for 30 min. (D and G) HEK293T cells transfected with NF-κB-luciferase reporter and indicated TLR7 mutants vectors and then treated with IMQ (10 µg/ml) for 24 h were harvested for luciferase assay. (H) Immunofluorescence staining of HEK293T cells transfected with GFP-TLR7 and TLR7 mutant vectors, prior to treatment of R848 (10 µg/ml) for 30 min, and labelled with specific antibodies. Scale bar: 100 μm. Data represent the mean ± SEM. *P* values are determined by two-tailed Student’s *t* test. \*\**P*<0.01, ns, not significant.

TLR7 has two predicted tyrosine residues (Y807, Y881) in the linker region (http://www.phosphosite.org/) and four tyrosine residues (Y891, Y897, Y972, Y1024) in the TIR domain. We thus constructed mutant plasmids of TLR7 by replacing tyrosine with glycine, and used them to transfected HEK293T cells with these plasmids. Western blots of endosomal proteins from HEK293T cells transfected with GFP-TLR7 or mutant vectors revealed that several mutations, viz. Y891A, Y897A, Y972A, and Y1024A, increased TLR7 expression relative to cells expressing wild-type TLR7 (Figure 6B). Immunoblotting assays showed that compared to wild-type TLR7, Y897A, Y972A, and Y1024A mutants enhanced TLR7 responses with p-p65 as a proxy (Figure 6C). NF-κB activation, assessed by a luciferase assay was also increased in the HEK293T cells overexpressing Y897A, Y972A, and Y1024A mutants of TLR7 (Figure 6D).

To further explore the effects of TLR7 phosphorylation on its downstream signaling, we constructed another set of mutant plasmids by replacing these tyrosine residues with aspartic acid to mimic lasting phosphorylation. Compared to HEK293T cells transfected with wild-type TLR7 vector, those transfected with respective GFP-TLR7 mutants expressed lower levels of TLR7, which was most obvious with Y1024D (Figure 6E). In terms of p65 phosphorylation and NF-κB activation, only Y1024D mutation caused marked attenuation (Figures 6F-6H).

### SHP2 mediates the ubiquitination of TLR7 via dephosphorylating its Y1024

We next explored the reason why phosphorylation of TLR7 affect downstream NF-*Κ*B activation. In Figure 5, SHP2 promotes the trafficking of TLR7 from Golgi to endosomes. While, ubiquitination plays a significant role in this trafficking. Therefore, we first demonstrated that TLR7 is ubiquitinated upon stimulation by TLR7 agonists, e.g., IMQ and R848 (Figure 7A). Next, we substitute two corresponding lysine residues in TLR7 (K951R and K952R), both of which reduced the phosphorylation levels of IKKα/β (Figure 7B), and not affect TLR7 phosphorylation (Figure 7C). To investigate which form of the polyubiquitin chain of TLR7 regulated by SHP2, we co-transfected GFP-TLR7 plasmid and HA-Ub or mutant Ubs and then immunoprecipitation with anti-GFP (Figure 7D), or only transfected GFP-TLR7 plasmid, and then used antibody to K63-linked ubiquitin (Figure 7E). Data showed that TLR7 polyubiquitination was K63-linked. Importantly, SHP2 also positively regulates TLR7 ubiquitination, as the ubiquitination level of TLR7 was obviously increased when SHP2 was over-expressed (Figure 7F), especially in an active form (Figure 7G). We then evaluated the effect of SHP2 on TLR7 ubiquitination, specifically through Y1024. In cells transfected with TLR7^Y1024A^ mutant mimicking de-phosphorylation, the ubiquitination of TLR7 was increased, similar to the effect of IMQ. However, in cells expressing the phospho-mimicking TLR7^Y1024D^, TLR7 was less ubiquitinated (Figure 7H). Furthermore, ubiquitination of wild-type TLR7 was increased by SHP2 overexpression to a level that was higher than when cells were transfected TLR7^Y1024D^ (Figure 7I). In line with these results, SHP2 knockdown using shRNA decreased the ubiquitination of wild-type TLR7, without affecting that of TLR7^Y1024A^ (Figure 7H). Similarly, SHP2 deficient HEK293T cells transfected with TLR7^Y1024A^, TLR7 ubiquitination almost unchanged (Figure 7J). Collectively, these lines of evidence suggest that SHP2 regulates the function of TLR7, specifically through dephosphorylation of TLR7, which promotes its ubiquitination and trafficking to endosomes.

**Figure 7.**
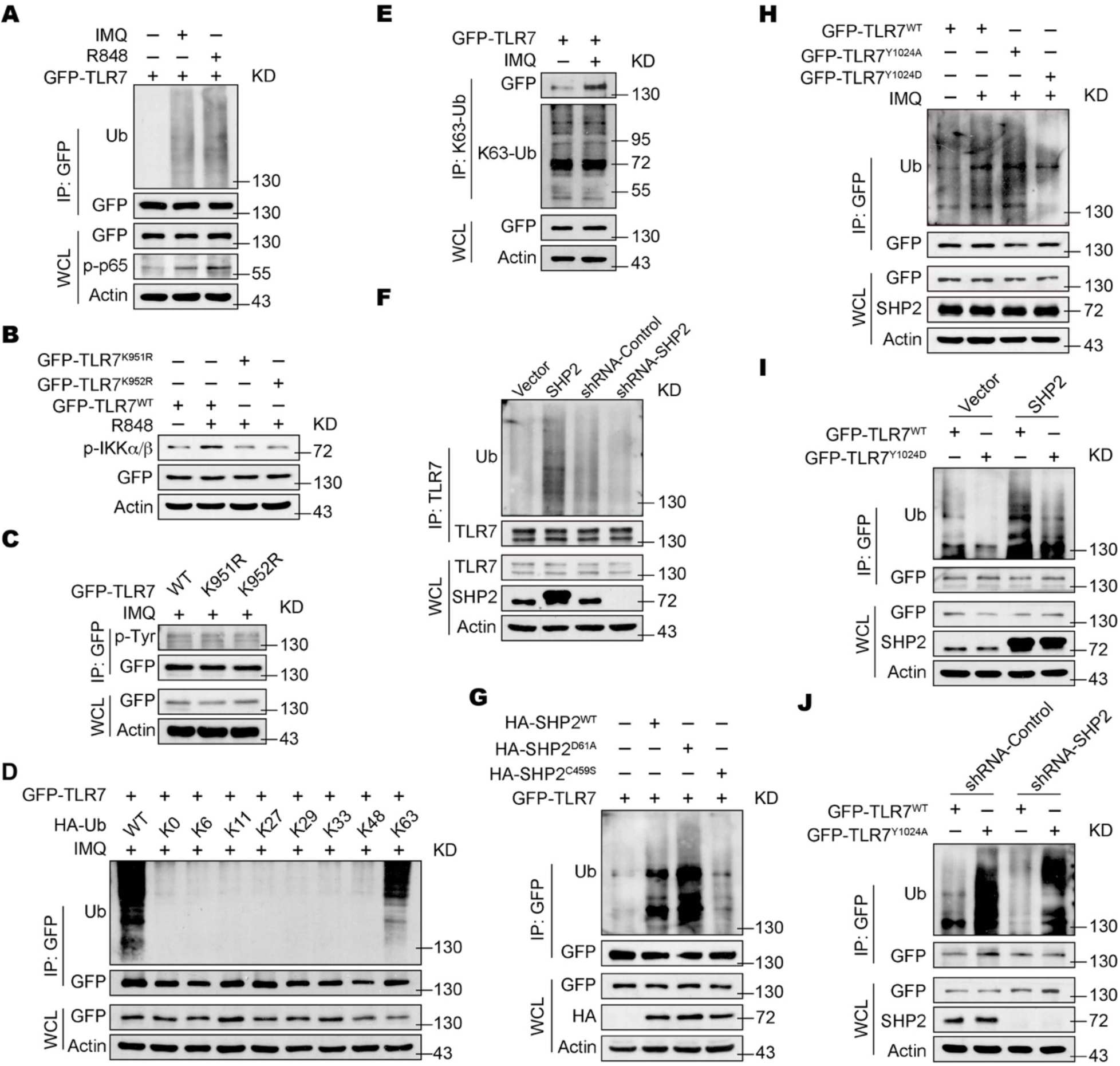
SHP2 mediates the ubiquitination of TLR7 via dephosphorylating its Y1024. (A) Immunoblotting of TLR7 ubiquitination in HEK293T cells transfected with GFP-TLR7 for 48 h, then treated with IMQ (10 μg/ml) or R848 (10 μg/ml) for 1 h. (B) Immunoblotting of HEK293T cells transfected with GFP-TLR7 and TLR7 mutant vectors and treated with or without R848 (10 µg/ml) for 30 min. (C) Immunoblotting of HEK293T cells transfected with GFP-TLR7 and TLR7 mutant vectors and treated with IMQ (10 µg/ml) for 30 min. (D) Immunoblotting of TLR7 ubiquitination in HEK293T cells co-transfected with GFP-TLR7, along with HA-Ub or mutant Ubs and treated with IMQ (10 µg/ml) for 1 h. (E) Immunoblotting of HEK293T cells transfected with GFP-TLR7 and treated with or without IMQ (10 µg/ml) for 30 min. (F) Immunoblotting of TLR7 ubiquitination in HEK293T cells with vector, SHP2, shRNA-Control or shRNA-SHP2 lentivirus were treated with IMQ (10 µg/ml) for 1 h. (G) HEK293T cells co-transfected with GFP-TLR7 and HA-SHP2 and HA-SHP2 mutant vectors for 48 h, and then infected with IMQ (10 µg/ml) for 1 h. (H) Immunoblotting of HEK293T cells transfected with GFP-TLR7 and TLR7 mutant vectors and treated with or without IMQ (10 µg/ml). (I) Immunoblotting of TLR7 ubiquitination in HEK293T cells with vector and SHP2 lentivirus transfected with GFP-TLR7 and GFP-TLR7^Y1024D^ plasmids and infected with IMQ (10 µg/ml) for 1 h. (J) Immunoblotting of HEK293T cells with shRNA-Control or shRNA-SHP2 lentivirus transfected with GFP-TLR7 and GFP-TLR7^Y1024A^ plasmids and infected with IMQ (10 µg/ml) for 1 h.

### SHP2 inhibitor attenuates the psoriasis-like phenotype in the IMQ-induced murine model

To determine whether SHP2 activity affects psoriasis pathogenesis, we treated the murine psoriatic model with SHP099, a potent allosteric inhibitor of SHP2 (Chen et al., 2016). SHP099 significantly suppressed IMQ-induced swelling, epidermal acanthosis, proliferation of keratinocytes, and dermal inflammatory cell infiltration (Figure 8A), attendant with a drastic decrease in the clinical scores (Figure 8B).

**Figure 8.**
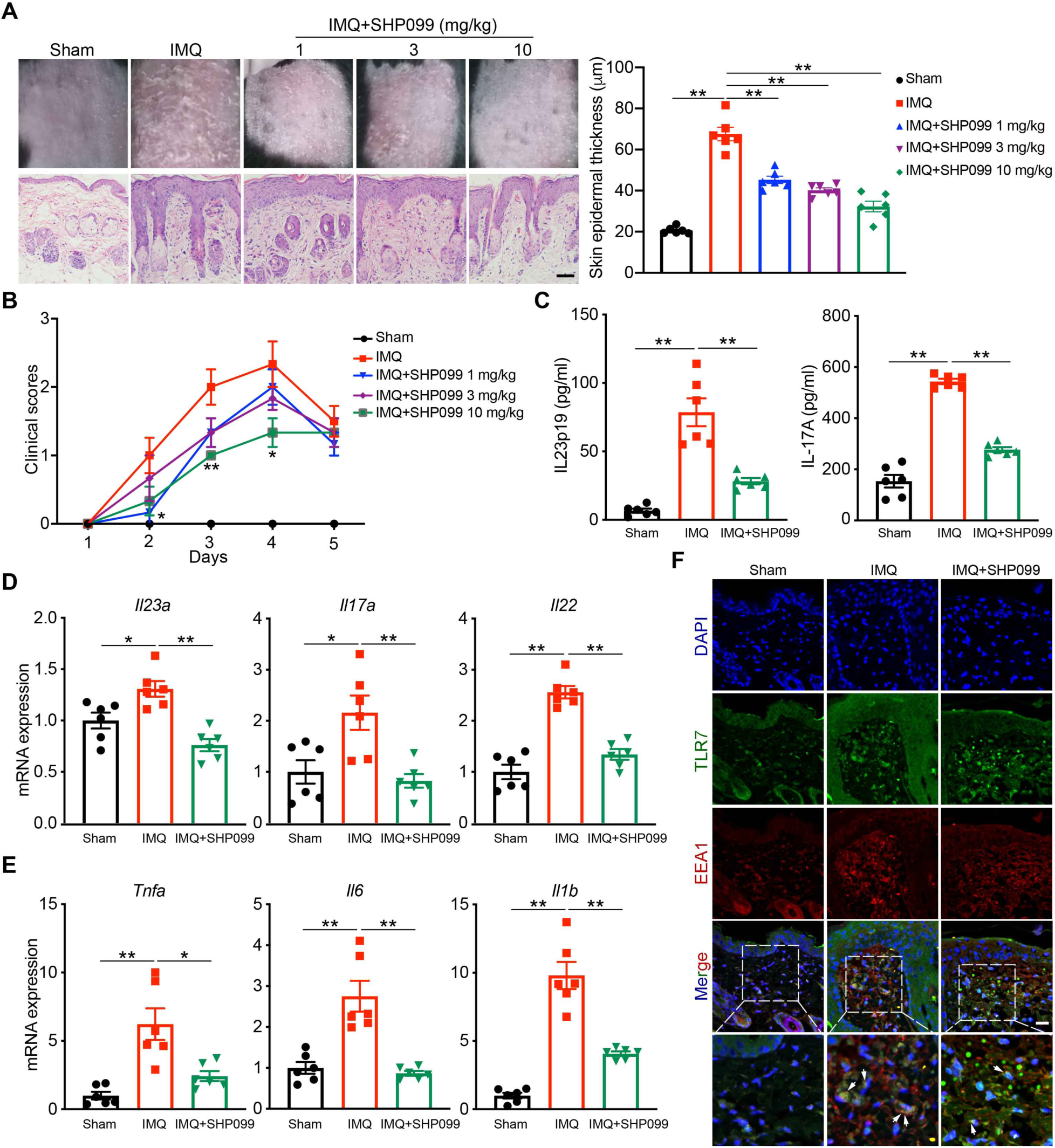
SHP2 inhibitor attenuates psoriasis-like phenotype in the IMQ-induced murine model. C57BL/6J mice (n = 6/group) were treated with indicated dose of SHP099 or vehicle for 4 days. (A) Phenotypic presentation (*top*) and H&E staining (*bottom*) of dorsal skin. Scale bar: 100 μm. Left: H&E staining data; right: statistical data (mean ± SEM). (B) Clinical scores plotted with mean ± SEM. * denotes statistical significance when compared with the IMQ group. (C) ELISA quantification of protein levels of cytokines in mouse serum. (D and E) Quantitative PCR analysis of mRNA encoding IL-23/IL-17A axis cytokines and other psoriasis-related cytokines (E) in the dorsal skin of C57BL/6J mice treated with indicated dose of SHP099 or vehicle for 4 days. Results were normalized to GAPDH expression. (F) Confocal microscopy imaging of skin sections from indicated mice labelled with anti-TLR7 (green), anti-EEA1 (red) and DAPI (blue), showing TLR7 expression on early endosome (arrows). Scale bar: 50 μm. Data represent mean ± SEM. *P* values are determined by two-tailed Student’s *t* test (A-E). \**P*<0.05, \*\**P*<0.01.

Consistent with the ameliorated psoriatic phenotype, the serum levels of IL-23 and IL-17A were also significantly reduced by SHP099 (Figure 8C). This was concordant with a decreased mRNA levels of genes related to the IL-23/IL-17 axis, such as *Il23a, Il17a*, and *Il22* (Figure 8D), as well as the expression of *Tnfa, Il6*, and *Il1b* in the skin (Figure 8E). RNA sequencing of peritoneal macrophages confirmed a significantly reduction of the mRNA expression of NF-κB signaling-related cytokines in response to IMQ stimulation *in vitro* due to SHP099 (Figure S9). SHP099 also significantly decreased TLR7 in endosomes, where it co-localized with EEA1 and LAMP1 (Figure 8F; Figure S10). Furthermore, SHP099 also markedly reduced the levels of NF-κB signaling-related cytokines in the murine model (Figures 8C-8E), as well as human PBMCs from healthy donors (Figure S11). Taken together, these data demonstrate that SHP099 ameliorates the IMQ-induced psoriatic development in mice by inhibiting TLR7-dependent NF-κB activation.

Finally, we tested the effect of SHP099 in the PBMCs derived from human donors. Expectedly, when compared to normal control donors, PBMCs obtained from psoriatic patients express significantly higher levels of NF-κB signaling-related cytokines. However, when these PBMCs were treated with 10 μM SHP099, the production of these cytokines was remarkably reduced, almost to a comparable level to that in the normal controls (Figure 9A). Collectively, our data demonstrate a positive regulation of psoriasis by SHP2. By dephosphorylating TLR7 at Tyr1024 and inducing TLR7 ubiquitination, SHP2 drives the trafficking of TLR7 to the endosome, which sustains the activation of downstream TLR7/NF-κB signaling, thereby promoting the development of psoriasis (Figure 9B).

**Figure 9.**
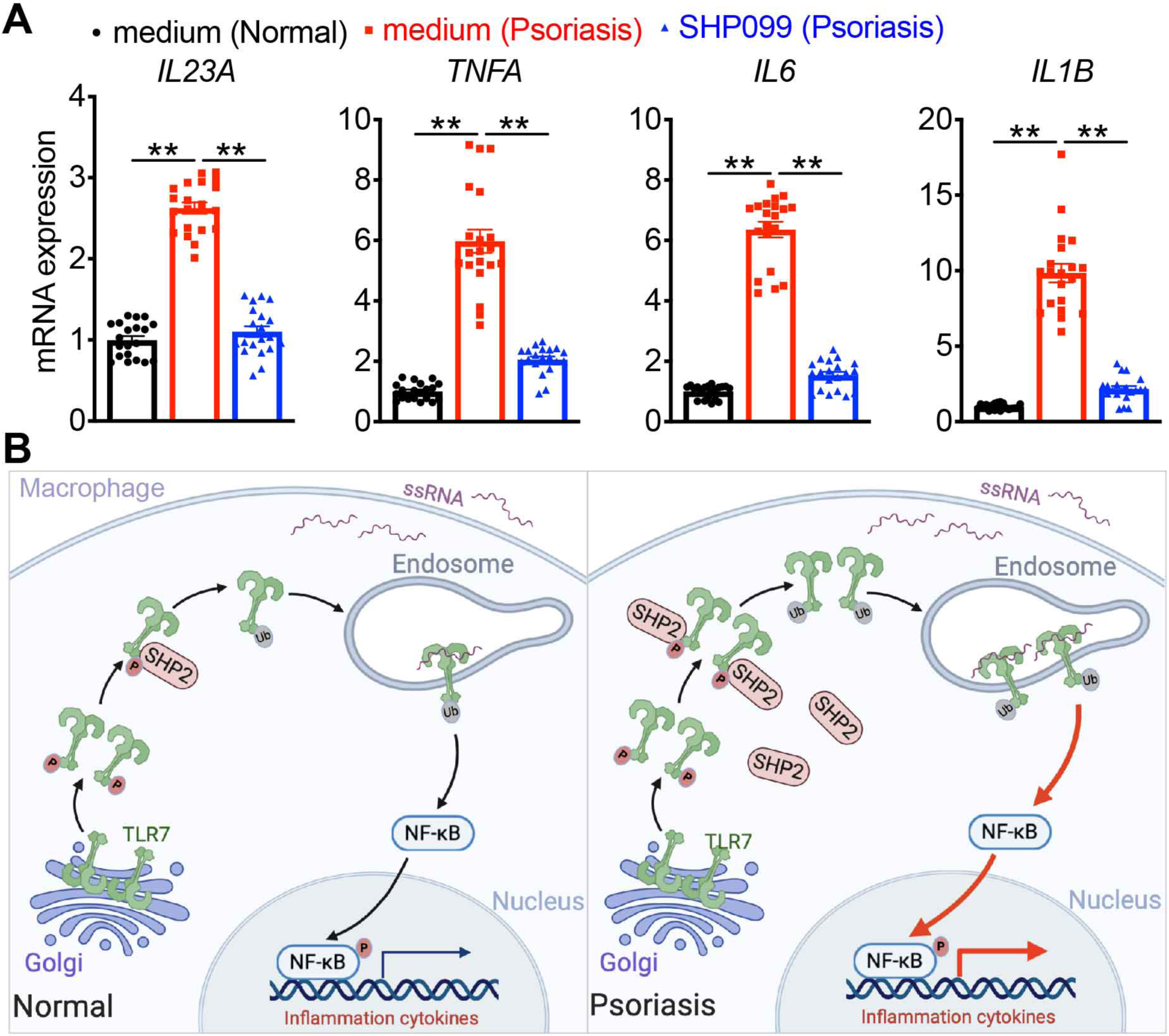
The graphic illustration of the mechanism of SHP2 accelerating the development of psoriasis. (A) Quantitative PCR analysis of mRNA levels in human PBMCs derived from psoriatic patients (n=20) and normal healthy controls (n=20). Data represent the mean ± SEM. *P* values are determined by two-tailed Student’s *t* test. \*\**P*<0.01. (B) In macrophages of normal skin, when stimulating by TLR7 agonist ssRNA (e.g., IMQ, R848), TLR7 is phosphorylated and SHP2 is recruited to dephosphorylate TLR7, then TLR7 is ubiquitinated, next activates downstream NF-κB signaling. However, in infiltrated macrophages of psoriatic skin, SHP2 is overexpressed. Hyper-dephosphorylation of TLR7 at Tyr1024 by SHP2 leads to hyper-ubiquitination of TLR7 and overactivation of NF-κB activation, resulting in unrestrained skin inflammation. Created in BioRender.com.

## Discussion

Accumulating evidence has revealed that SHP2 is closely related to autoimmune disease (Wang et al., 2016). Herein, we found that in infiltrated macrophages of psoriatic skin, SHP2 expression is highly increased. As a consequence, SHP2-mediated TLR7/NF-κB activation is augmented, leading to further increases in the expressions of psoriasis-related inflammatory cytokines including IL-23A, TNF-α, IL-6 and IL-1β, and accelerates psoriasis-like skin inflammation. SHP2 deficiency in macrophages neither in dendritic cells nor in γδT cells (Kadekar et al., 2020) or SHP2 allosteric inhibitor attenuates IMQ-induced psoriasis-like phenotype. Together, our results demonstrate that SHP2 is a critical regulator of psoriatic development and suggest SHP2 as a potential therapeutic target for the treatment of psoriasis.

Both the innate and adaptive immune systems have been considered as important drivers of psoriasis. Recently, the IL-23/IL-17 axis has been shown to play a pivotal role in psoriasis (Boehncke and Schön, 2015; Burkett and Kuchroo, 2016; Kopp et al., 2015; Lebwohl, 2019), and several negative regulators of this axis have been identified, including adaptor proteins, microRNAs, and etc. Mechanisms mediated by these regulators in T cells, macrophages, and keratinocytes have been identified to attribute to the development of psoriasis (Bambouskova et al., 2018; Boisson et al., 2013; Srivastava et al., 2017; Wang et al., 2018; Wu et al., 2018; Xu et al., 2018; Yan et al., 2015). These include ACT1, a critical adaptor protein in the IL-17 signaling pathway. A missense single nucleotide polymorphism (rs33980500; SNP-D10N) in the coding region of ACT1 was suggested to increase the compensatory Th17 cells, with excessive secretion of IL-22 and IL-17, constituting a main cause of psoriasis susceptibility in ACT1-D10N patients (Boisson et al., 2013). Another negative regulator is microRNA (miR)-146a, the deficiency of which contributes to IL-17-driven inflammation in keratinocytes, early disease onset, and aggravated skin inflammation in IMQ-induced model (Srivastava et al., 2017). On the other hand, a number of other molecules, e.g. microRNA-210 (Wu et al., 2018) and IκBζ (Bambouskova et al., 2018), IL-25 (Xu et al., 2018), Card14 (Wang et al., 2018) and NF-κB (Yan et al., 2015) have been identified as positive regulators of psoriasis, through regulating various processes, including Th17 cell differentiation, IL-17-driven inflammation, and proliferation of keratinocytes (Bambouskova et al., 2018; Wang et al., 2018; Wu et al., 2018; Xu et al., 2018; Yan et al., 2015). In addition, antimicrobial peptide LL37 has been shown to enable keratinocytes to produce IFN-β, which promotes maturation of dendritic cells and contributes to the pathogenesis of psoriasis (Zhang et al., 2016). The extensive findings from these previous studies strongly point to a central role of NF-kB activation and IL-23/IL-17 signaling in psoriasis. Here, our data show that in infiltrated macrophages of psoriatic skin, SHP2 is strongly induced and acts as a positive regulator. Via dephosphorylation of TLR7 specifically at Tyr1024, SHP2 triggers a sequela of molecular events, including 1) ubiquitination of TLR7, 2) the trafficking of TLR7 to the endosome, and 3) excessive activation of TLR7/NF-κB signaling, fueling the uncontrolled inflammation. Ablation or inhibition of SHP2 in macrophages reduces the production of pro-inflammatory cytokines and prevents the development of psoriasis-like skin inflammation in both the imiquimod-induced murine model and psoriatic patients. Taken together, our findings connecting SHP2 and TLR7 highlight the importance of SHP2 as a positive regulator of inflammation in psoriasis and likely in other autoimmune diseases.

Toll-like receptors (TLRs) are essential players in innate and adaptive immunity. Intracellular TLRs, such as TLR3, TLR7, TLR8 and TLR9, are intrinsically capable of recognizing nucleic acids to induce immune responses (Petes et al., 2017). In particular, TLR7 recognizes ssRNA and acts via MyD88, activating two major signaling pathways including NF-κB and IFN-regulatory factors (IRFs) to trigger the transcription of genes encoding proinflammatory cytokine and the production of type I IFNs (Hu et al., 2016). Previous studies have shown that TLR7 activation leads to upregulation of proinflammatory cytokines to accelerate the development of psoriasis (Kim et al., 2018). TLR7 overexpression, as a result of TLR8 deficiency in DCs leads to augmented NF-κB activation in response to TLR7 ligands and thereby aggravates the development of spontaneous autoimmunity (Demaria et al., 2010). Interestingly, *Datura metel* L., a traditional Chinese medicine shown to suppress the progression of IMQ-induced psoriasis in the mouse model, can suppress TLR7 and TLR8 expression and inhibit the activity of TLR7/8-MyD88-NF-κB-NLRP3 inflammasome pathway (Yang et al., 2019). Our data demonstrate that SHP2 can promote the localization of TLR7 to the endosome, without affecting TLR7 expression. SHP2 overexpression or overactivation (e.g. that observed in IMQ or psoriasis) maintains excessive activation of TLR7/NF-κB signaling and accelerates psoriasis-like skin inflammation. Mechanistically, we found that SHP2 drives TLR7 translocation in a phosphatase-dependent manner, specifically by dephosphorylating TLR7 at Tyr1024. Although several TLRs including TLR2, TLR3, TLR4, TLR5, TLR8 and TLR9 can be tyrosine phosphorylated upon stimulation in the cytoplasmic TIR domain (Chattopadhyay and Sen, 2014), it is not known whether and how TLR7 can be (de)phosphorylated at the tyrosine residues. To the best of our knowledge, this study is the first report showing that TLR7 can be phosphorylated at Tyr1024 and its dephosphorylation by SHP2 accelerates TLR7 ubiquitination, promotes TLR7 activation of NF-kB signaling, which contributes to the aggravation of psoriasis.

SHP2 is a ubiquitously expressed tyrosine phosphatase encoded by *PTPN11*. It was the first cloned phosphatase containing SH2 domain (Feng et al., 1993). SHP2, as an intracellular signaling molecule responding to various cytokines, growth factors and other extracellular stimulators, is ubiquitously expressed in various cells of the body, and participates in various aspects of cellular physiology including cell proliferation, activation, migration and differentiation (Tajan et al., 2015). We have previously demonstrated that upon treatment of NLRP3 inflammasome stimulators, SHP2 translocates to mitochondria where it interacts with mitochondrial endomembrane protein adenine nucleotide translocase 1 (ANT1) and dephosphorylates ANT1 to maintain mitochondrial homeostasis. This serves as a mechanism to negatively regulate the activation of NLRP3 inflammasome (Guo et al., 2017). In this context, SHP2 is regarded as an anti-inflammatory factor. Other studies suggest that SHP2 can also act as a negative regulator of inflammation. For example, SHP2 has been identified as a detrimental factor for inflammatory bowel disease (IBD) by disrupting macrophage response to interleukin-10 (Xiao et al., 2019). On the other hand, a study performed in the context of systemic lupus erythematosus (SLE) showed that SHP2 was increased in PBMCs isolated from patients with the SLE compare to the normal controls. A SHP2 inhibitor 11a-1 reversed the symptoms of SLE-associated organ damage, suggesting that SHP2 is a positive regulator development of systemic lupus erythematosus (Wang et al., 2016). In the current study, we found that the levels of SHP2 in PBMCs and skin lesions of psoriatic patients are significantly higher than those in normal controls. Using macrophage-specific conditionally SHP2 knockout mice (SHP2^lyz2-/-^) or a SHP2 allosteric inhibitor SHP099, we demonstrated that SHP2 inhibition attenuated IMQ-induced psoriasis-like phenotype, suggesting that SHP2 is a positive regulator in the psoriatic disease progression. Collectively, the pro- or anti-inflammatory role of SHP2 in inflammatory diseases is context-dependent, which involves the intricate balance among various immune cells and numerous chemokines and cytokines.

In summary, we unraveled a novel regulation by SHP2 in TLR7-mediated NF-κB signaling activation in the macrophages. Such positive regulation of SHP2, through dephosphorylating TLR7 at Tyr1024, promotes the trafficking of TLR7 to the endosome, maintains excessive activation of TLR7/NF-κB signaling. Together, our studies identify SHP2 as a novel driver for pathogenesis of psoriasis and suggest that SHP2 inhibition as a promising therapeutic approach for the treatment of autoimmune disease, such as psoriasis.

## Data Availability

The RNA-seq data have been deposited to the Gene Expression Omnibus with accession no. GSE147535. All other data needed to evaluate the conclusions in this manuscript are present in the paper.

## Acknowledgments

We thank Prof. Cliff Y. Yang (Sun Yat-sen University, China) for providing Itgax-Cre mice. We thank Dr. Lifei Hou (Harvard Medical School, Boston, MA) for valuable advice on this project.

## Methods

### Study design

The goal of this study was to explore the role of SHP2 in the development of psoriasis. To assess this, we analyzed the mRNA and protein expression level of SHP2 in human PBMCs from healthy donors and psoriatic patients. We also used SHP2 conditional knockout mice to explore it. The mice were randomly assigned to different groups. For all studies, animal numbers were depicted in the figure legends. To point out the therapeutic potential of selective SHP2 inhibition in IMQ-induced psoriasis-like mouse model, we used SHP2 allosteric inhibitor to prove it. Primary data were located in Data files.

### Mice

C57BL/6J mice were purchased from GemPharmatech Co. Ltd. The macrophage-specific SHP2 knockout mice (SHP2^lyz2-/-^) were generated by crossing SHP2^flox/flox^ mice with Lyz2-Cre transgenic mice. The dendritic cell-specific SHP2 knockout mice (SHP2^Itgax-/-^) were generated by crossing SHP2^flox/flox^ mice with Itgax-Cre transgenic mice. These mice were bred and maintained in specific pathogen-free conditions at GemPharmatech Co. Ltd and Experimental Animal Center at Nanjing University. Age- and sex-matched mice 6-10 weeks of age were used. Animal welfare and experimental procedures were approved by the Institutional Animal Care and Use Committee at Nanjing University. All efforts were made to reduce the number of animals used and to minimize animal suffering.

### Human specimens

All human studies were approved by the Ethics Institutional Review Board of West China Hospital, Sichuan University. Normal human skin was obtained from patients at West China Hospital undergoing elective surgeries in which skin was discarded as a routine part of the procedure. Psoriatic skin biopsies and blood were obtained from psoriatic patients with active clinical disease (study number 2019-R-513). PBMCs were isolated from the peripheral blood of psoriatic patients and normal healthy donors. Written informed consents were obtained from all patients.

### SHP2 enzyme activity assay

The enzyme activity of SHP2 in PBMCs was measured using the surrogate substrate DiFMUP in a fluorescence assay. The phosphatase reactions were performed in 96-well black plate (Corning) at room temperature. The final reaction volume was 100 μL including 60 mM HEPES, pH 7.2, 75 mM NaCl, 75 mM KCl, 1 mM EDTA, 0.05% P-20, 5 mM DTT. PBMCs of normal controls and psoriasis patients were lysed in Western and IP lysis buffer (Beyotime) supplemented with protease (MCE). Proteins were quantified by the Bradford assay (HyClone-Pierce). There are two groups: proteins (1 μg) were incubated with PHPS1 (an PTP inhibitor) (Sigma-Aldrich) at 25 °C for 30 min and added the surrogate substrate DiFMUP (Invitrogen) and incubated at 25 °C for 30 min. The reaction was quenched by the addition of bpV (Phen) (Enzo Life Sciences). The fluorescence signal was measured using a microplate reader (Envision) using excitation and emission wavelengths of 340 and 450 nm, respectively. Another group is the same except without PHPS1. Finally, the subtraction of these two groups is the enzyme activity of SHP2 in PBMCs.

### IMQ-induced psoriasis-like mouse model

Eight- to ten-week-old female C57BL/6J and SHP2^lyz2-/-^ mice were used for establishing the IMQ-induced psoriasis-like model. The mice were treated with a daily topical dose of 62.5 mg of IMQ cream (5%) (Aldara; 3M Pharmaceuticals) on the shaved dorsal skin for 4 consecutive days. Mice were sacrificed 1 day later after the last treatment. For each mouse, skin lesions were taken for hematoxylin and eosin (H&E) staining, immunocytochemistry, immunofluorescence and quantitative real-time PCR analysis. In addition, serum was also collected for cytokine assays by ELISA.

### Histological analysis

Harvested mouse dorsal skin and human skin tissues were flushed with PBS, fixed with 4% formaldehyde overnight and embedded in paraffin. Sections (5 μm thick) were stained with hematoxylin and eosin (H&E) according to standard procedures.

For immunohistochemistry, the human and mouse skin paraffin sections were deparaffinized, rehydrated, and antibody retrieved with sodium citrate, blocked, then stained with anti-SHP2 (Santa Cruz, catalog sc-7384), anti-ERK (Cell Signal Technology, catalog 4695), anti-CD68 (Cell Signal Technology, catalog 76437), anti-p-p65 (Cell Signal Technology, catalog 3033), anti-Ki67 (Abcam, catalog ab15580) were used at 1:100 overnight at 4°C. After 3 rinses of 1 × PBS, the slides were detected using Real Envision Detection kit (GeneTech) according to the manufacturer’s instructions.

For immunofluorescence, paraffin-embedded human and mouse skin sections were deparaffinized and rehydrated, antigen retrieved with sodium citrate, blocked with 5% goat serum, and incubated with primary Abs overnight at 4°C. Anti-SHP2 (Santa Cruz, catalog sc-7384), anti-CD68 (Cell Signal Technology, catalog 76437), anti-p-p65 (Cell Signal Technology, catalog 3033), anti-TLR7 (Novus, catalog NBP2-24906), anti-EEA1 (Santa Cruz, catalog sc-137130), anti-LAMP1 (Santa Cruz, catalog sc-20011), anti-Rab11a (Santa Cruz, catalog sc-166912), anti-LAT1 (Santa Cruz, catalog sc-53550), anti-Calreticulin (Abcam, catalog ab92516), anti-Giantin (Abcam, catalog ab80864), anti-TLR7 (Santa Cruz, catalog sc-57463) were used in 1:100. After 3 rinses of 1 × PBST, sections were treated with Alexa Fluor 633-conjugated donkey anti-goat IgG (H+L) cross-adsorbed secondary Ab (1:500; Invitrogen, catalog A-21082), Alexa Fluor 488-conjugated goat anti-rabbit IgG (H+L) cross-adsorbed secondary Ab (1:500; Invitrogen, catalog A-11034), Alexa Fluor 488-conjugated goat anti-mouse IgG (H+L) cross-adsorbed secondary Ab (1:500; Invitrogen, catalog A-32723), Alexa Fluor 546-conjugated goat anti-mouse IgG (H+L) cross-adsorbed secondary Ab (1:500; Invitrogen, catalog A-11003), Alexa Fluor 594-conjugated goat anti-rabbit IgG (H+L) cross-adsorbed secondary Ab (1:500; Invitrogen, catalog A-11037) at room temperature for 2 hours in the dark, and the nuclei were stained with DAPI. All the cells were imaged by an inverted confocal microscope (Carl Zeiss).

### Immunocytochemistry

PMA (Sigma-Aldrich)-differentiated THP-1 cells or PMs were incubated in 24-well culture dishes at a density of 1 × 10^5^ cells per well. Cells were washed by PBS and then fixed by 4% paraformaldehyde for 10 minutes at room temperature. Following 3 rinses in 1 × PBS, cells were permeabilized using 0.5% Triton X-100 (Beyotime) for 30 minutes at 4°C. After blocking cells with 5% BSA for 1 h, cells were cultured with primary Abs overnight at 4°C. Anti-TLR7 (Novus, catalog NBP2-24906), anti-SHP2 (Santa Cruz, catalog sc-7384), anti-EEA1 (Santa Cruz, catalog sc-137130), anti-LAMP1 (Santa Cruz, catalog sc-20011), anti-EEA1 (R&D systems, catalog AF-8047) were used at 1:100. After 3 rinses of 1 × PBST, coverslips were treated with Alexa Fluor 633-conjugated donkey anti-goat IgG (H+L) cross-adsorbed secondary Ab (1:500; Invitrogen, catalog A-21082), Alexa Fluor 488-conjugated goat anti-rabbit IgG (H+L) cross-adsorbed secondary Ab (1:500; Invitrogen, catalog A-11034), Alexa Fluor 488-conjugated goat anti-mouse IgG (H+L) cross-adsorbed secondary Ab (1:500; Invitrogen, catalog A-32723), Alexa Fluor 546-conjugated goat anti-mouse IgG (H+L) cross-adsorbed secondary Ab (1:500; Invitrogen, catalog A-11003), Alexa Fluor 594-conjugated goat anti-rabbit IgG (H+L) cross-adsorbed secondary Ab (1:500; Invitrogen, catalog A-11037) at room temperature for 2 hours in the dark, and the nuclei were stained with DAPI. All the cells were imaged by an inverted confocal microscope (Carl Zeiss).

### Quantitative PCR (qPCR)

Total RNA was extracted from the skin tissues of the mice, BMDMs, PMs or THP-1 cells using TRIzol (TaKaRa) as described by the manufacturer. Single-stranded cDNA was synthesized from 1 μg of total RNA by reverse transcription. Quantitative PCR was performed on a CFX 100 (Bio-Rad, Hercules, CA) cycler using the primers listed in Supplemental Table 1. The amplification program was as follows: 95°C for 2.5 min, and 44 cycles at 95°C for 15 s, 60°C for 30 s. Dissociation curves were analyzed at the end of the amplification. The level of GAPDH or Actin RNA expression was used to normalize the data.

### RNA-seq analysis

Peritoneal macrophages from wild-type and SHP2^lyz2-/-^ mice (8-10-week old) were stimulated with IMQ 4 h later, PMs were used for total RNA isolation with Trizol (TaKaRa), and subjected to RNA-seq analysis. RNA sequencing libraries were generated using NEBNext® Ultra™ RNA Library Prep Kit for Illumina® (NEB, USA) following manufacturer’s recommendations. Raw data (raw reads) of fastq format were firstly processed through in-house perl scripts. Index of the reference genome was built using Hisat2 v2.0.4 and pairedend clean reads were aligned to the reference genome using Hisat2 v2.0.4. HTSeq v0.9.1 was used to count the numbers of reads mapped to each gene. Differential expression analysis was performed using the DESeq R package. The *P* values were adjusted using the Benjamini & Hochberg method. Corrected *P*-value of 0.005 and log 2 (fold change) of 1 were set as the cut-off for significantly differential expression. The accession number for the RNA-seq reported in this paper in GEO is GSE147535.

### Isolation of peritoneal macrophages

Mice were intraperitoneally injected with 1 ml starch broth per mouse. After 3 days, peritoneal cells were harvested by lavaging the peritoneal cavity with 5 ml PBS. Floating cells were removed by DMEM washing and adherent peritoneal macrophages were cultured in DMEM containing 10% FBS and 1X penicillin/streptomycin at 37°C overnight, followed by treatment with different stimuli according to experimental designs.

### Generation of bone marrow-derived macrophages

Mice were sacrificed by cervical dislocation and removal of both femurs and tibia. Bone marrow cells were isolated by flushing with DMEM. Red blood cells were lysed using NH4Cl. Bone marrow cells were cultured in DMEM containing 10% FBS supplemented with M-CSF (10 ng/ml) for 5-7 days. Two days later, non-adherent cells were removed, with fresh media replenished, and cultured for two more days. BMDMs were used on day 5-7.

### Immunoprecipitation

First, HEK293T cells were transfected with various epitope-tagged vectors. After 48 hours incubation, we extracted protein using lysis buffer supplemented with protease and phosphatase inhibitors. Each sample was incubated with anti-GFP or anti-HA antibodies overnight at 4°C. Then incubated with magnetic beads (Millipore) at room temperature for 1 hour. Proteins not immobilized on beads were removed by five times washes with cold lysis buffer. Next, the precipitates were boiled for 10 min in SDS loading buffer. The presence or absence of the target protein, was evaluated by Western blot using the indicated antibodies.

### Western blotting

Cells or skin samples were lysed in radio immunoprecipitation assay (RIPA) buffer supplemented with protease and phosphatase inhibitor (MCE). Proteins were quantified by the Bradford assay (HyClone-Pierce). The proteins were then separated by SDS-polyacrylamide gel electrophoresis (PAGE) and electrophoretically transferred onto polyvinylidene difluoride membranes. The membranes were probed with antibodies overnight at 4°C, and then incubated with a horseradish peroxidase-coupled secondary antibody. Detection was performed using a LumiGLO chemiluminescent substrate system. Anti-SHP2 (1:1,000; Santa Cruz, catalog sc-7384), anti-p-IKK*α*/*β* (1:1,000; Cell Signal Technology, catalog 2697), anti-IKK*α* (1:1,000; Cell Signal Technology, catalog 11930), anti-IKK*β* (1:1,000; Cell Signal Technology, catalog 8943), anti-p-p65 (1:1,000; Cell Signal Technology, catalog 3033), anti-p65 (1:1,000; Cell Signal Technology, catalog 8243), anti-TLR7 (1:1,000; Abcam, catalog ab113524), anti-TLR7 (1:1000; Santa Cruz, catalog sc-57463), anti-TLR7 (1:1000; Novus, catalog NBP2-24906), anti-MyD88 (1:1,000; Cell Signal Technology, catalog 4283), anti-NaK-ATPase (1:1,000; Cell Signal Technology, catalog 23565), anti-Calreticulin (1:1,000; Abcam, catalog ab92516), anti-Giantin (1:1,000; Abcam, catalog ab80864), anti-EEA1 (1:1,000; Cell Signal Technology, catalog 3288), anti-Calnexin (1:1,000; Abcam, catalog ab92573), anti GFP-tag (1:1,000; Santa Cruz, catalog sc-9996) and anti-HA-tag (1:1,000; Cell Signal Technology, catalog 3724), anti-A20 (1:1,000; Cell Signal Technology, catalog 5630), anti-TRAF6 (1:1,000; Cell Signal Technology, catalog 8028), anti-p-Tyr (1:1,000; Santa Cruz, catalog sc-7020), anti-Ub (1:1000; Santa Cruz, catalog sc-8017), anti-β-actin (1:2,000, Abmart, catalog M20011), anti-β-tubulin (1:2,000, Abmart, catalog M20005) were used.

### Flow cytometry

Single-cell suspensions from BMDMs and THP1 cells were subjected to flow cytometry using Attune NxT (Thermo Fisher) and the following fluorescence-labeled antibodies from Invitrogen: PE-conjugated anti-TLR7 (MA5-16247); FITC-conjugated anti-TLR7 (PA5-23127). All FACS data were analyzed by FlowJo 10.4.

## Isolation of Membrane, ER, Golgi, and Endosome Protein

### Membrane

Membrane Protein Extraction Kit (Thermo Scientific) was used to isolate membrane protein according to the manufacturer’s instruction. Briefly, 5 × 10^6^ cells were collected and washed in cold PBS. After centrifugation, added Permeabilization Buffer to the cell pellet and centrifuged. Added Solubilization Buffer to the pellet and incubated at 4°C for 30 minutes. After centrifugation, transferred supernatant containing solubilized membrane and membrane-associated proteins to a fresh tube.

### ER

ER Enrichment Kit (Invent Biotechnologies) was used to isolate ER according to the manufacturer’s instruction. Briefly, 3 × 10^7^ cells were collected and added buffer A. Then the cell suspension was transferred to a filter cartridge and were centrifuged twice. Next, transferred all supernatant to a fresh tube and centrifuged. After centrifugation, transferred supernatant to a fresh tube and added buffer B and centrifuged. Resuspended the pellet cold buffer A and vortexed vigorously. Added buffer C and vortexed briefly. Transferred supernatant to a fresh tube and added buffer D. After centrifugation, the pellet was dissolved in buffer WA-009.

### Golgi

Golgi Apparatus Enrichment Kit (Invent Biotechnologies) was used to isolate Golgi according to the manufacturer’s instruction. Briefly, 3 × 10^7^ cells were collected and washed in cold PBS, then transferred to a filter cartridge. After a few times centrifuged, transferred supernatant to a fresh tube and added equal buffer B and vortexed briefly. After centrifugation, the pellet was resuspended in cold buffer A and centrifuged. Transferred supernatant to a fresh tube and added cold buffer C and vortexed vigorously. After centrifugation, the pellet was dissolved in buffer WA-009.

### Endosome

Endosome Isolation and Cell Fractionation kit (Invent Biotechnologies) was used to isolate endosome according to the manufacturer’s instruction. Briefly, 3 × 10^7^ cells were collected and added buffer A. Then the cell suspension was transferred to a filter cartridge and were centrifuged. The pellet was resuspended and centrifuged, next transferred the supernatant to a fresh tube and added buffer B and were incubated at 4°C overnight. After centrifugation, the pellet was dissolved in buffer WA-009.

### Luciferase assay

1 × 10^5^ HEK293T cells were seeded in the wells of a 24-well plate overnight, and each well was co-transfected with 500 ng different TLR7 mutant vectors, 100 ng of the firefly luciferase reporter plasmid pGL3 containing NF-*Κ*B promoter together with 0.5 ng of *β*-gal-promoter-dependent Renilla luciferase reporter. After 24 h transfection, cells were lysed. Firefly and Renilla luciferase activities were measured with the Dual-Luciferase Reporter System (Promega), and Renilla luciferase activity was used to normalize for transfection efficiency.

### ELISA

ELISA assays were used to detect for the presence of various cytokines in mouse serum isolated from IMQ-induced murine model animals. ELISA kits for mouse cytokines IL-23A and IL-17A were purchased from eBioscience. ELISA detections were all performed according to the manufacturer’s instructions.

### Statistics

Statistical analysis was performed using GraphPad Prism 8.0. Data are presented as the mean ± SEM. We assessed data for normal distribution and similar variance between groups. Statistical significance (\**P* < 0.05, \*\**P* < 0.01) was assessed using two-tailed unpaired Student’s *t* test for comparisons between 2 groups. When the data were not normally distributed or displayed unequal variances between 2 groups, we used the two-tailed Mann-Whitney U test for statistical analysis. No animals were excluded from statistical analysis.

## Supplementary Figures

**Figure S1.**
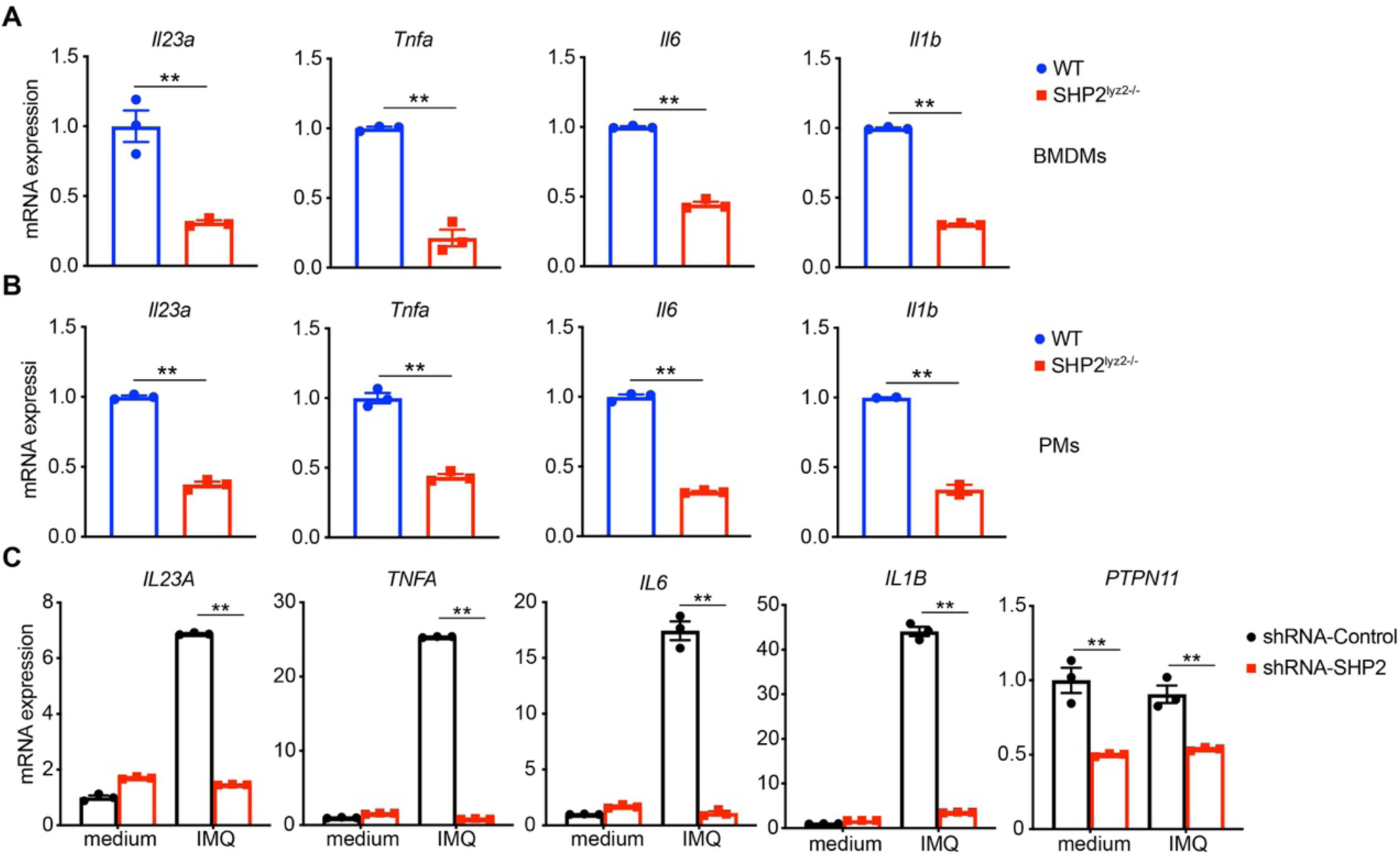
SHP2-deficient macrophages produce less psoriasis-related cytokines with IMQ stimulation. (A and B) Quantitative PCR analysis of mRNA levels in the (A) bone-marrow derived macrophages (BMDMs) and (B) peritoneal macrophages (PMs) derived from wild-type and SHP2^lyz2-/-^ mice treated with IMQ (10 µg/ml). Results were normalized to glyceraldehyde-3-phpsphate dehydrogenase (GAPDH) expression and are shown as mean ± SEM. (C) Quantitative PCR analysis of the indicated genes using PMA-differentiated THP-1 cells with shRNA-Control or shRNA-SHP2 lentivirus stimulated with IMQ (10 µg/ml) for 6 h. mRNA levels were normalized relative to β-actin. Data represent the mean ± SEM. *P* values are determined by two-tailed Student’s *t* test. \*\**P*<0.01.

**Figure S2.**
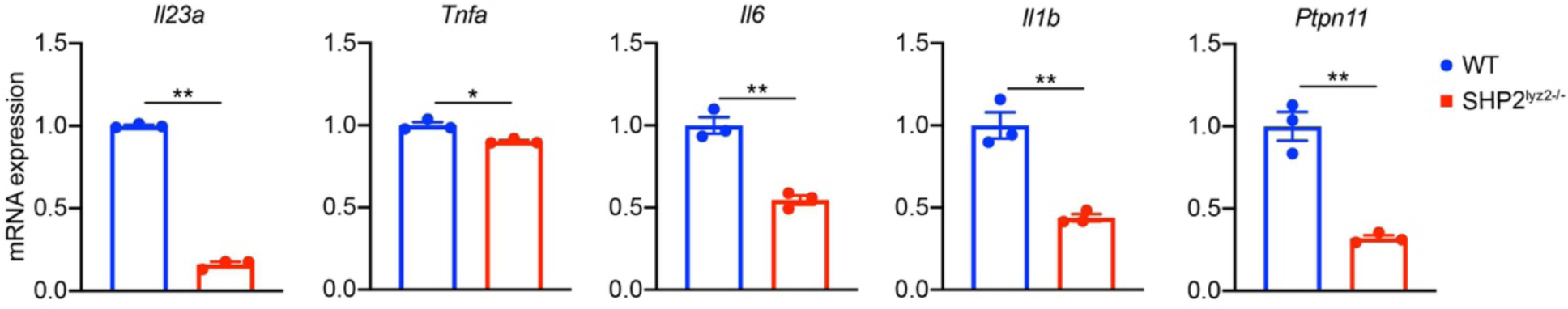
SHP2-deficient peritoneal macrophages reduce the levels of psoriasis-related cytokines with IL-36 stimulation. Expression levels of representative psoriasis-related genes of peritoneal macrophages derived from wild-type and SHP2^lyz2- /-^ mice treated with IL-36 (100 ng/ml) for 6 h. Data represent the mean ± SEM. *P* values are determined by two-tailed Student’s *t* test. \**P*<0.05, \*\**P*<0.01.

**Figure S3.**
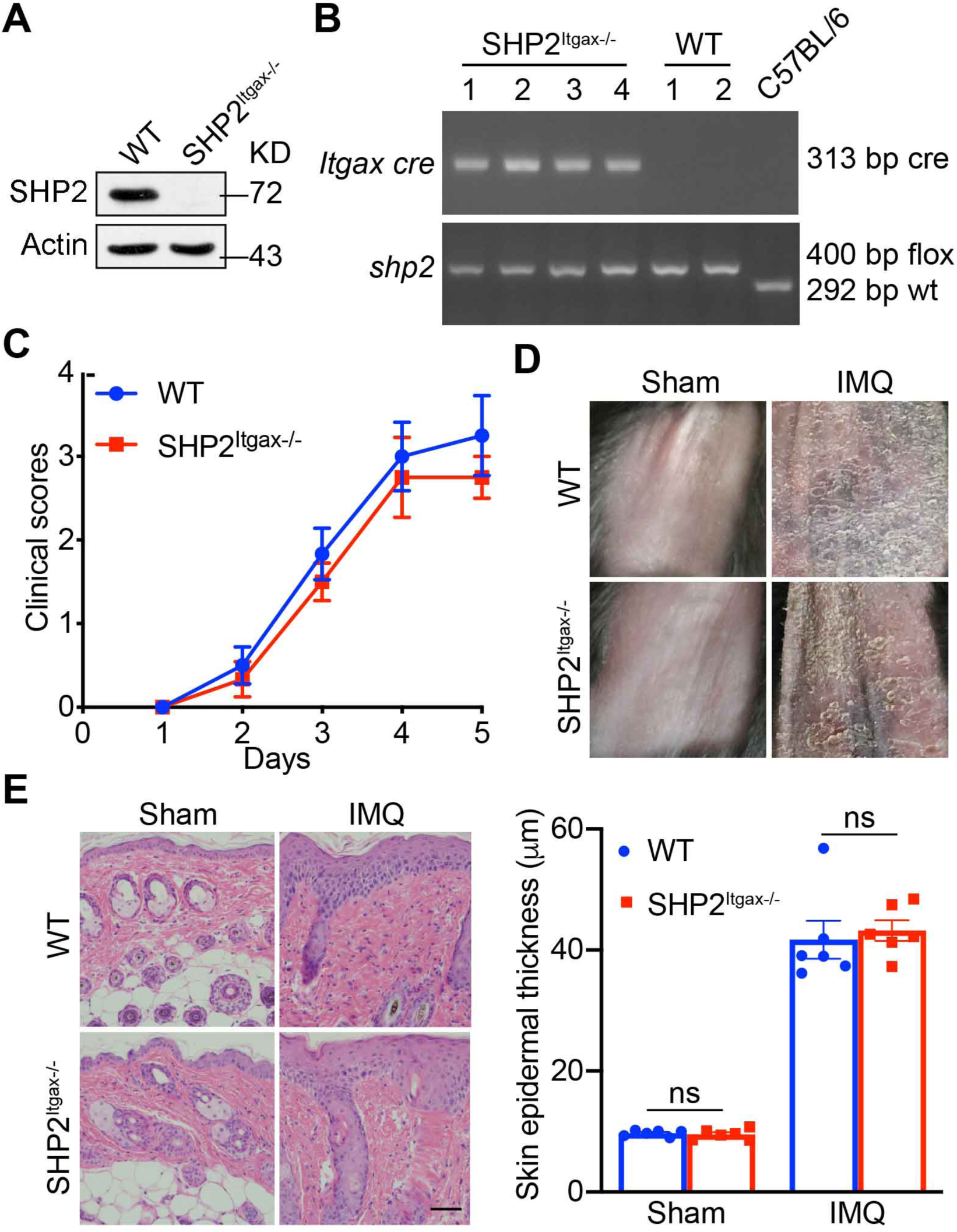
SHP2 deficiency in dendritic cells cannot alleviate psoriasis-like phenotype in the IMQ-induced murine model. (A and B) SHP2 protein level (A)and mRNA expression level (B) in BMDCs of wild-type and SHP2^Itgax-/-^ mice (n = 6/group). (C) Clinical scores or (D) phenotypic presentation of wild-type and SHP2^Itgax-/-^ mice treated with or without IMQ for 4 days. (E) H&E staining of mouse back skin of wild-type and SHP2^Itgax-/-^ mice treated with or without IMQ for 4 days. Left: H&E staining data; right: statistical data (means ± SEM). Scale bar: 100 μm. *P* values are determined by two-tailed Student’s *t* test. ns, not significant.

**Figure S4.**
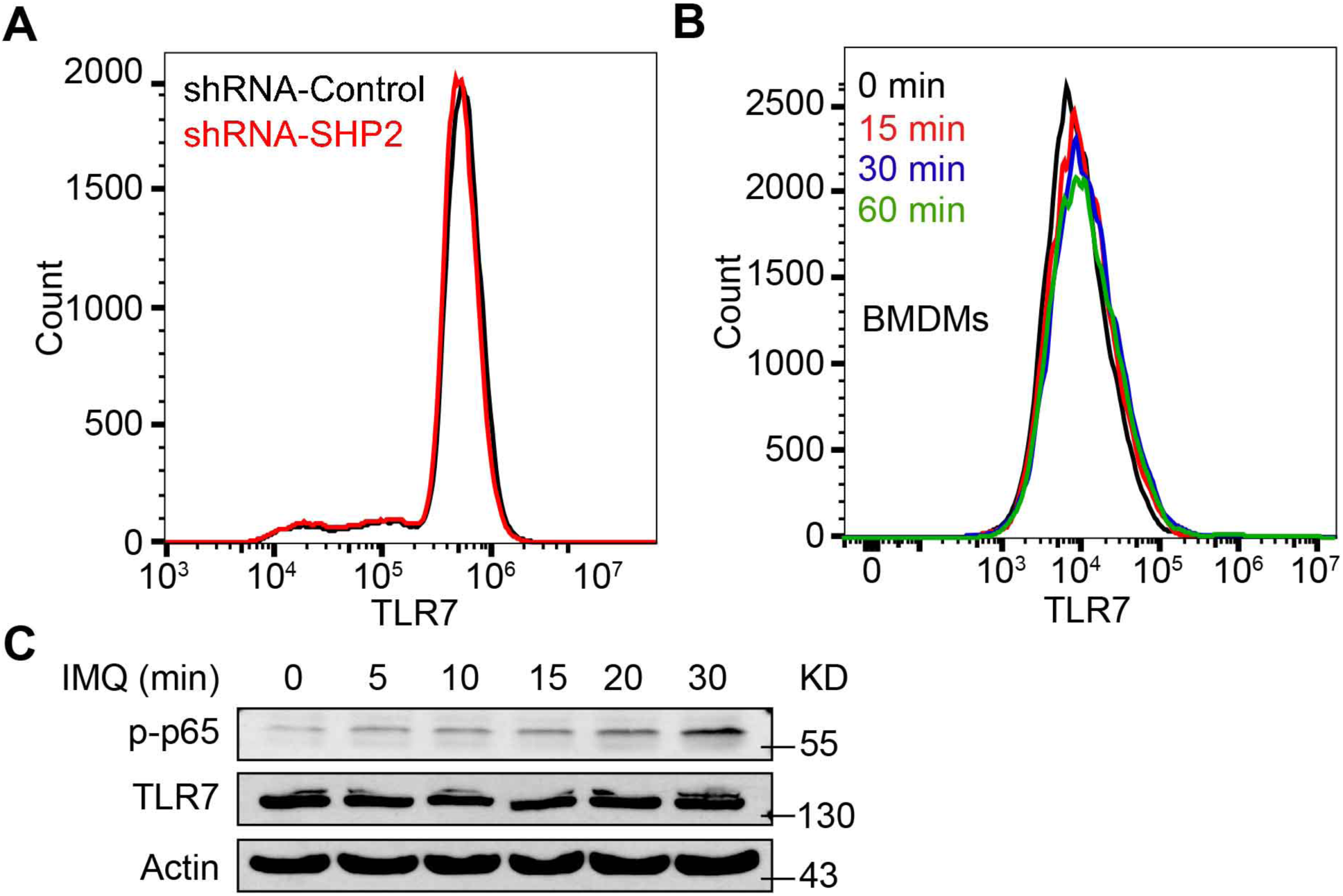
SHP2 unchanges protein level of TLR7. (A) Flow cytometry analysis of TLR7 expression on bone-marrow derived macrophages (BMDMs) from C57BL/6J mice treated by IMQ (10 µg/ml) for various times as indicated. (B) Flow cytometric analyses of TLR7 expression on PMA-differentiated THP-1 cells with shRNA-Control or shRNA-SHP2 lentivirus. (C) PMA-differentiated THP-1 cells were either untreated or treated by IMQ (10 µg/ml) for various times as indicated. Cell lysates were immunoblotted by respective antibodies.

**Fig S5.**
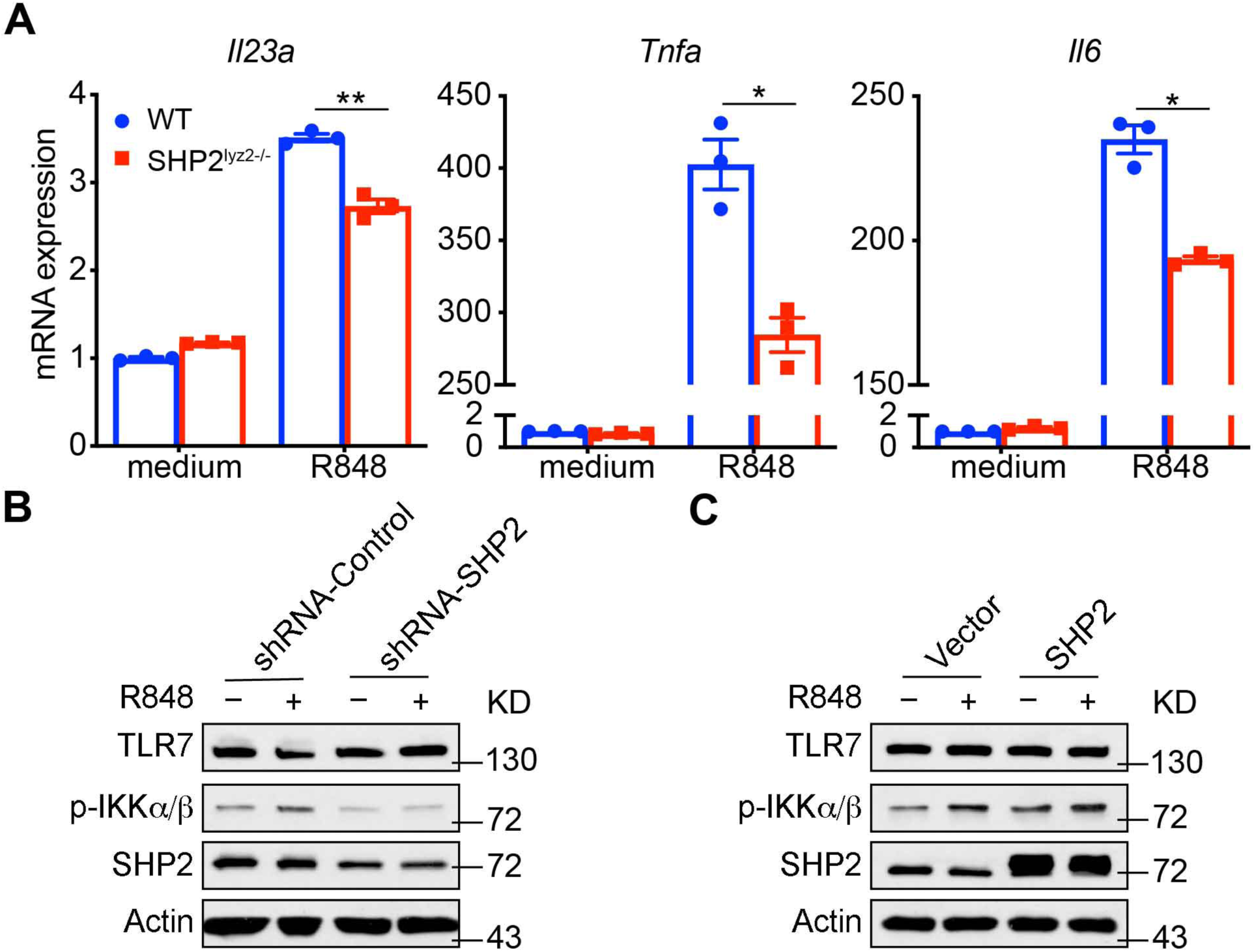
SHP2 deficiency reduces levels of psoriasis-related cytokines in R848-induced macrophages. (A) Expression levels of representative psoriasis-related genes of peritoneal macrophages derived from wild-type and SHP2^lyz2-/-^ mice treated with R848 (10 µg/ml) for 6 h. (B and C) PMA-differentiated THP-1 cells with shRNA-Control, shRNA-SHP2 (B), vector or SHP2 lentivirus (C) were unstimulated or stimulated by R848 (10 µg/ml) for 30 min. Whole cell lysates were resolved by 10% SDS-PAGE and then probed by respective antibodies. Data represent the mean ± SEM. *P* values are determined by two-tailed Student’s *t* test. \**P*<0.05, \*\**P*<0.01.

**Figure S6.**
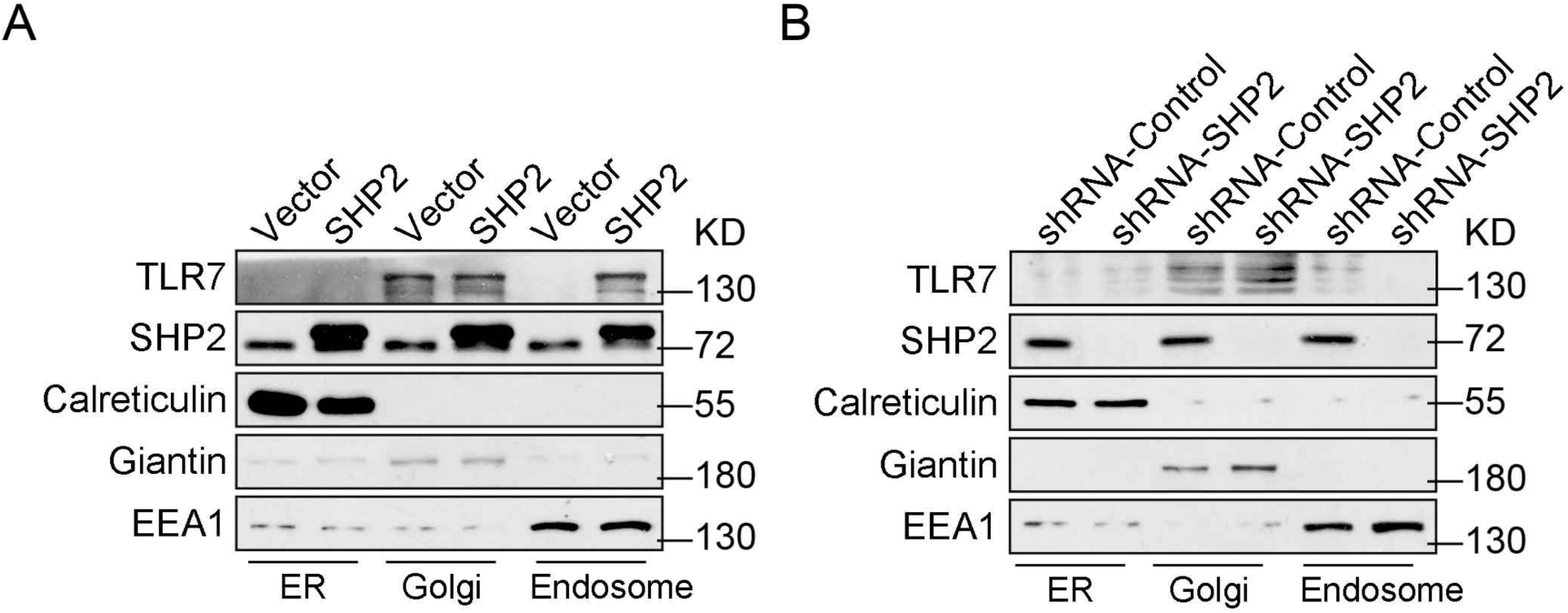
SHP2 affects the localization of TLR7 in endosome. (A and B) Immunoblot analysis of TLR7 or SHP2 in the isolated ER, Golgi and endosome from PMA-differentiated SHP2 overexpressed (A) and SHP2 deficient (B) HEK293T cells. **Figure S7**

**Figure S7.**
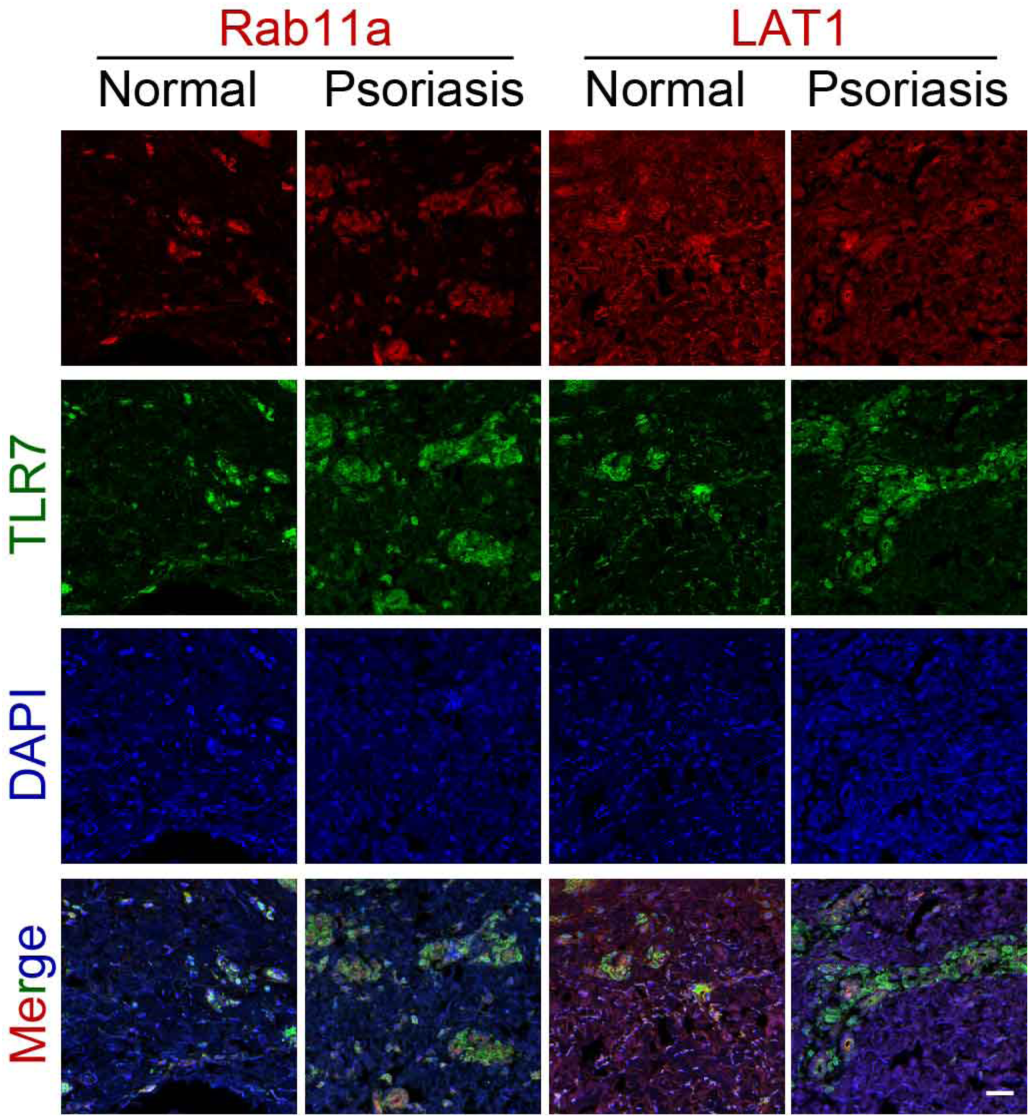
Cellular localization of TLR7 in psoriatic skin. Skin sections from psoriasis patients and healthy controls were immunostained for TLR7 together with a marker of the recycling endosome (Rab11a) or cell membrane (LAT1) prior to analysis by confocal microscopy. Scale bar: 100 μm.

**Figure S8.**
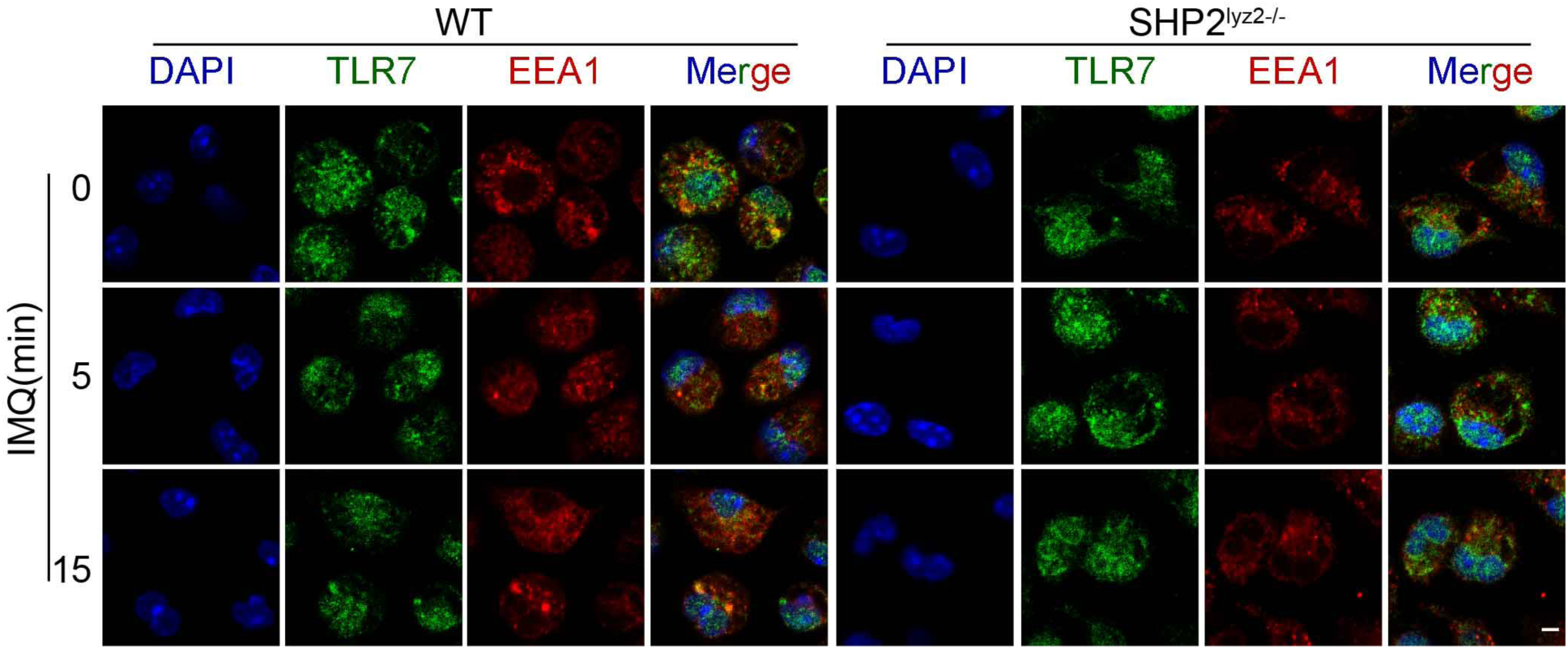
TLR7 is decreased in endosome of SHP2-deficient peritoneal macrophages. Confocal microscopy imaging of peritoneal macrophages derived from wild-type and SHP2^lyz2-/-^ mice were infected for various times as indicated with IMQ (10 µg/ml) and labelled with antibodies to the appropriate protein. Scale bar: 20 µm.

**Figure S9.**
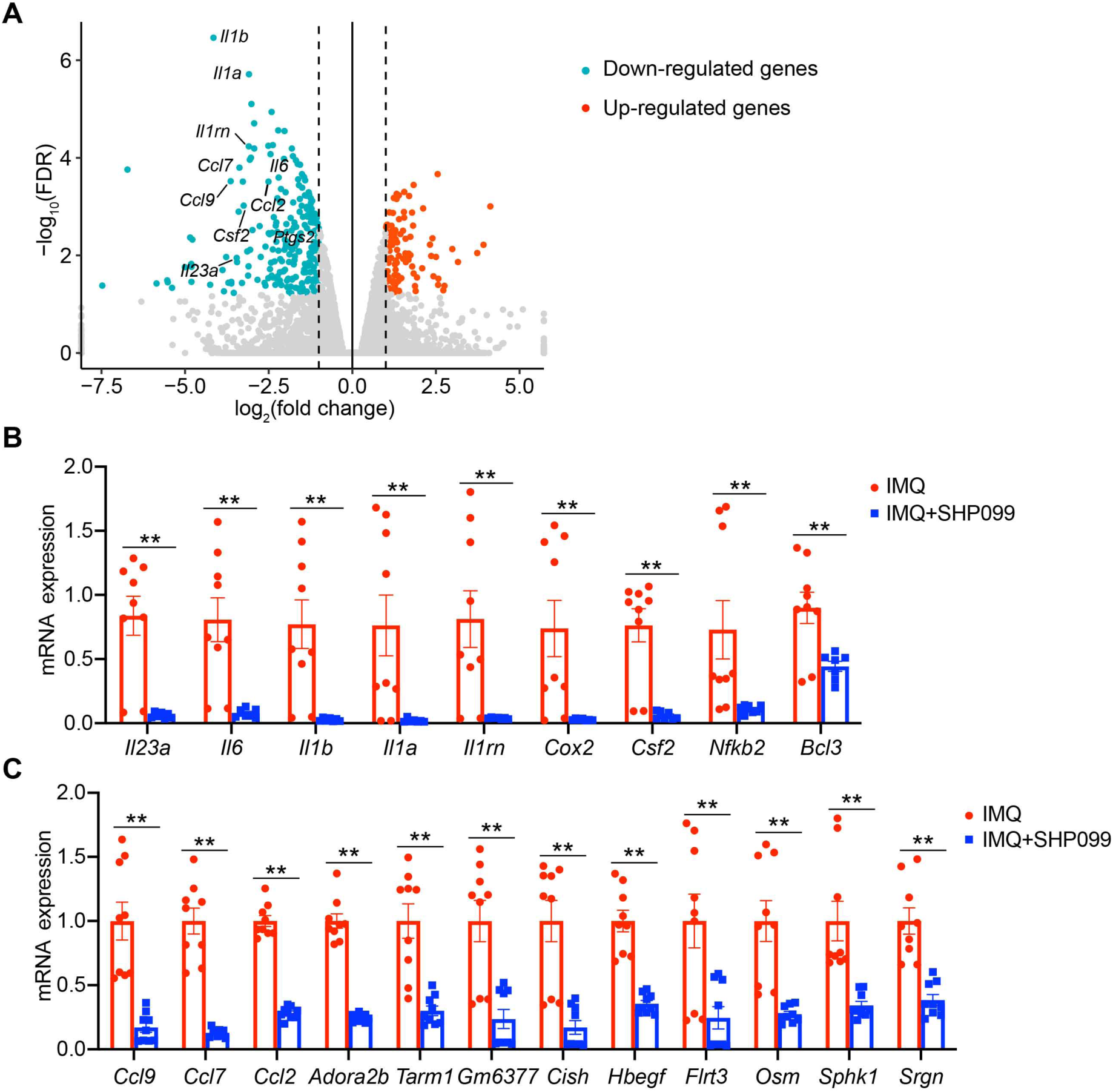
SHP2 allosteric inhibitor SHP099 prevents psoriasis by down-regulating NF-DB activation. (A) Volcano plot image of up-regulated genes (red) and down-regulated genes (green) from peritoneal macrophages derived from C57BL/6J mice untreated or pre-treated with SHP099 (10 µM) for 2 h and then stimulated by IMQ (10 µg/ml) for 4 h (n=3/group). (B and C) Peritoneal macrophages derived from C57BL/6J mice were untreated or pre-treated with SHP099 (10 µM) for 2 h and then stimulated by IMQ (10 µg/ml) for 4 h. Expression levels of indicated genes decreased in the SHP099 group compared to medium control.

**Figure S10.**
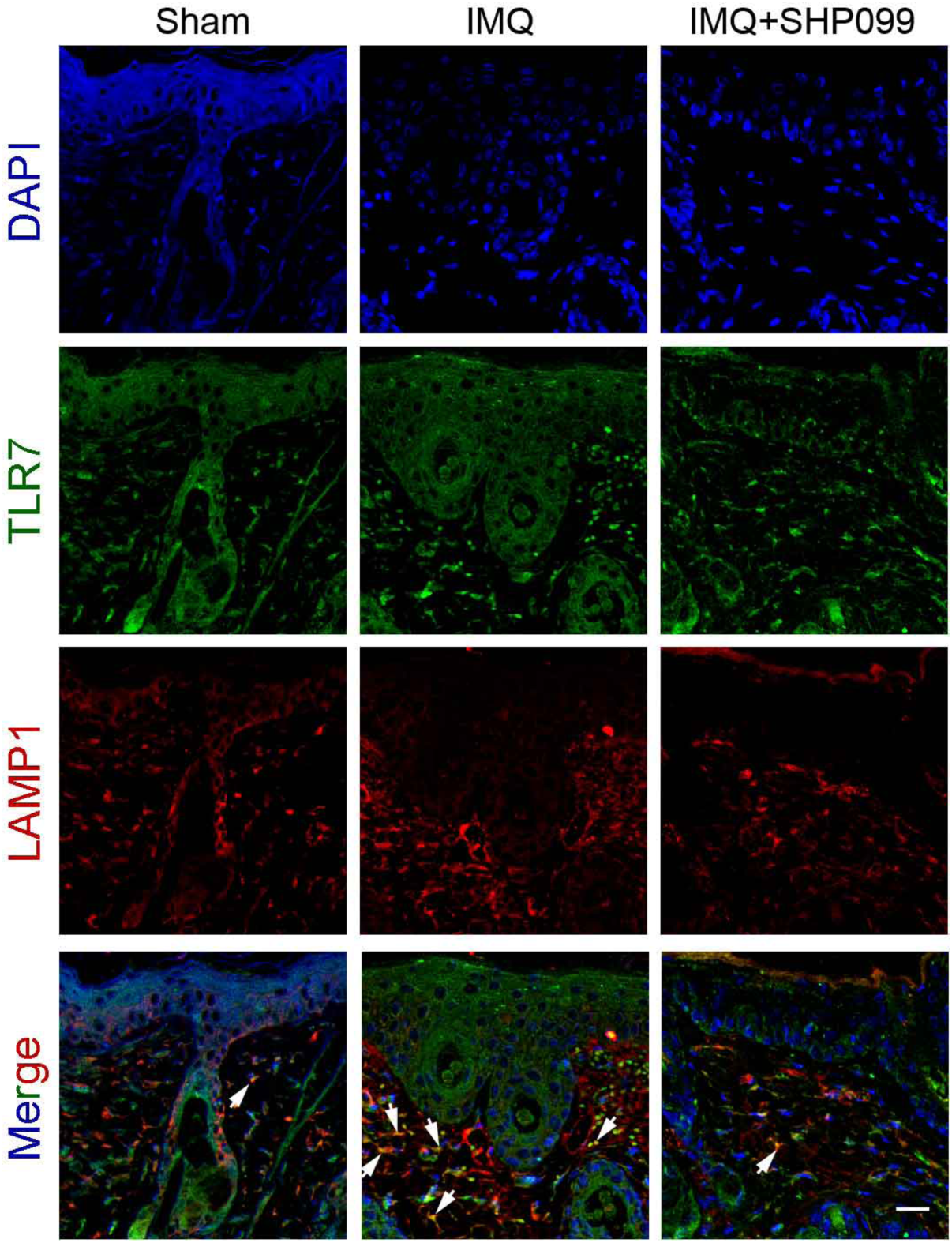
The localization of TLR7 in endosome is decreased in SHP099-treated group. Confocal microscopy imaging of skin sections from indicated mice labelled with anti-TLR7 (green), anti-LAMP1 (red) and DAPI (blue), showing TLR7 expression on lately endosome (arrows). Scale bar: 50 μm.

**Figure S11.**
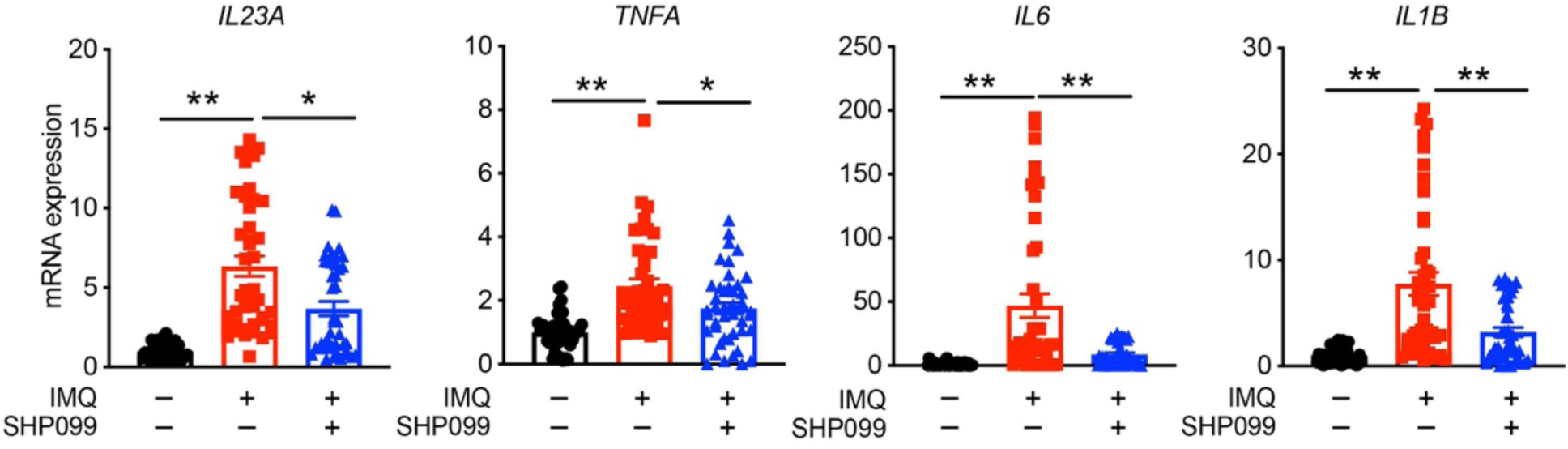
SHP2 inhibitor reduces levels of psoriasis-related cytokines in human peripheral blood mononuclear cells. Quantitative PCR analysis of mRNA levels of human peripheral blood mononuclear cells derived from healthy donors (n=43) were untreated or pre-treated with SHP099 (10 µM) for 2 h and then stimulated by IMQ (10 µg/ml) for 6 h. The mRNA levels were normalized relative to β-actin.

**Table S1.**
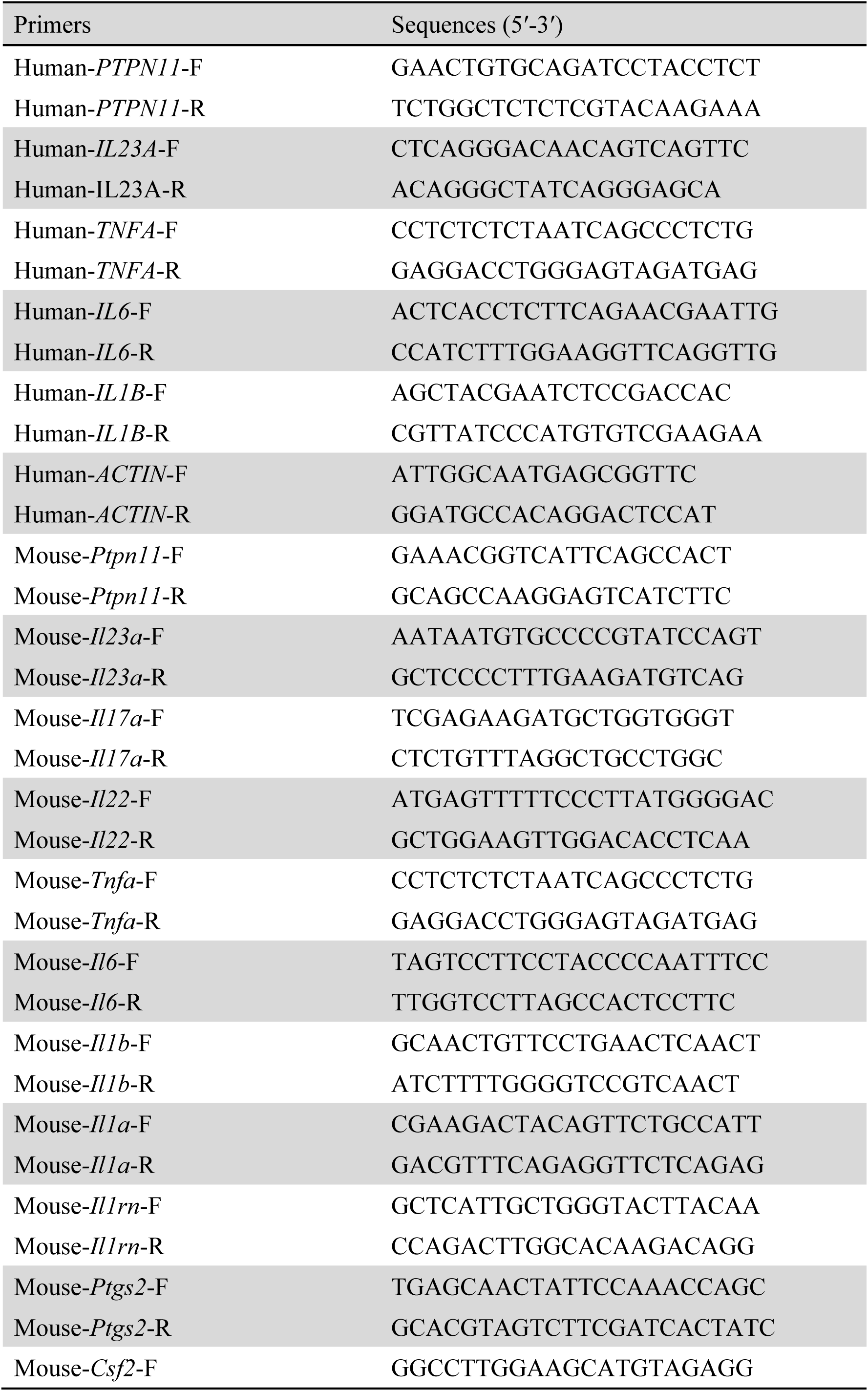
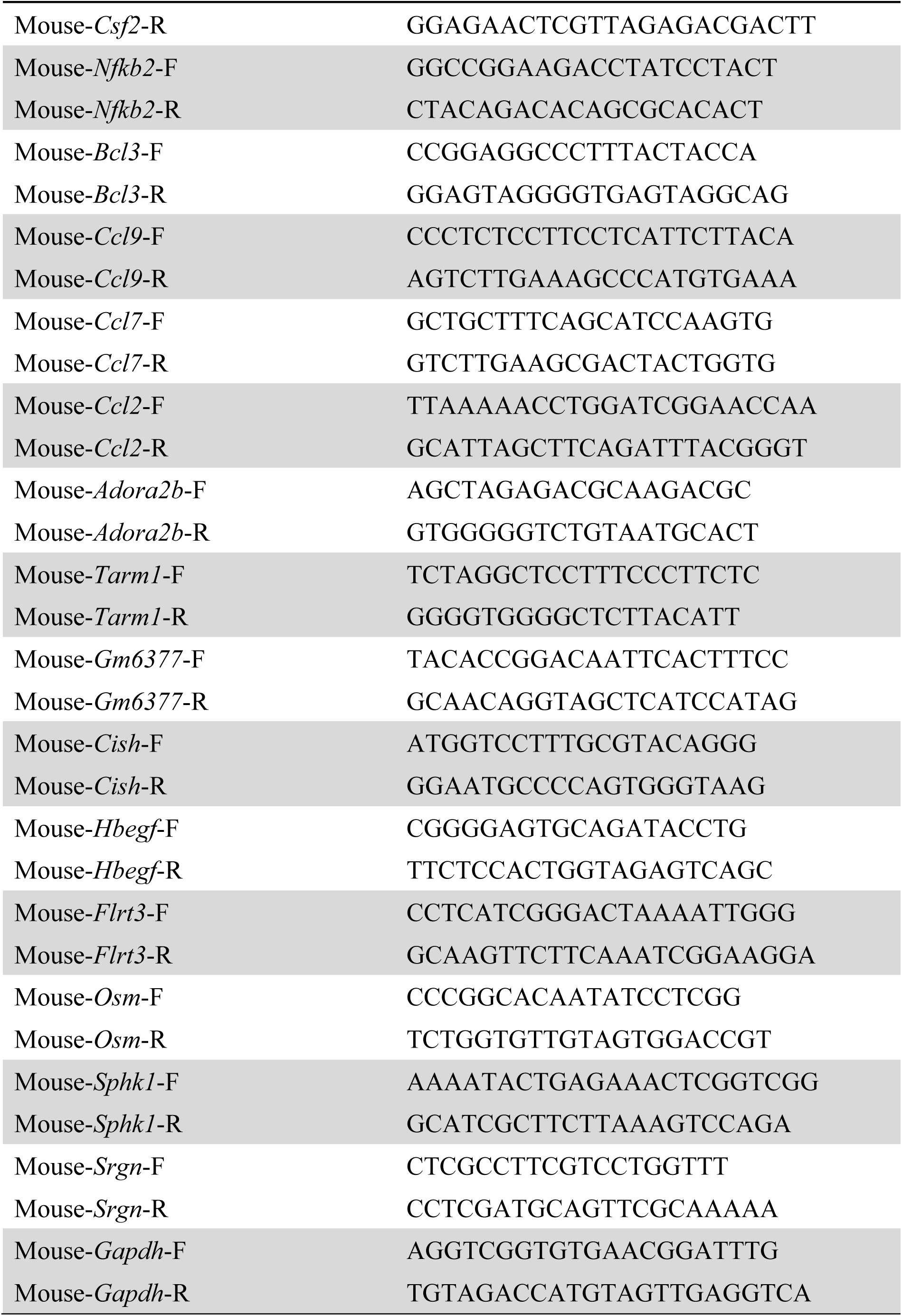
Primers for quantitative PCR analysis.

